# Widespread environment-specific causal effects detected in the UK Biobank

**DOI:** 10.1101/2024.08.21.24312360

**Authors:** Leona Knüsel, Alice Man, Guillaume Paré, Zoltán Kutalik

## Abstract

**Background:** Mendelian Randomization (MR) is a widely used tool to infer causal relationships. Yet, little research has been conducted on the elucidation of environment specific causal effects, despite mounting evidence for the relevance of causal effect modifying environmental variables.

**Methods:** To investigate potential modifications of causal effects, we extended two-stage-least-squares MR to investigate interaction effects (2SLS-I). We first tested 2SLS-I in a wide range of realistic simulation settings including quadratic and environment-dependent causal effects. Next, we applied 2SLS-I to investigate how environmental variables such as age, socioeconomic deprivation, and smoking modulate causal effects between a range of epidemiologically relevant exposure (such as systolic blood pressure, education, and body fat percentage) - outcome (e.g. forced expiratory volume (FEV1), CRP, and LDL cholesterol) pairs (in up to 337’392 individuals of the UK biobank).

**Results:** In simulations, 2SLS-I yielded unbiased interaction estimates, even in presence of non-linear causal effects. Applied to real data, 2SLS-I allowed for the detection of 182 interactions (P<0.001), with age, socioeconomic deprivation, and smoking being identified as important modifiers of many clinically relevant causal effects. For example, the positive causal effect of Triglycerides on systolic blood pressure was significantly attenuated in the elderly whilst the positive causal effect of Gamma-glutamyl transferase on CRP was intensified in smokers.

**Conclusion:** We present 2SLS-I, a method to simultaneously investigate environment-specific and non-linear causal effects. Our results highlight the importance of environmental variables in modifying well-established causal effects.

## Introduction

### Causal Effects and the role of Mendelian Randomization

The distinction between associations and causal relationships is a central challenge in epidemiology, as understanding causal effects – in contrast to mere associations - allows for the design of effective interventions.

To characterize causal effects, randomized controlled trials (RCTs) are considered as the gold standard. In RCTs, individuals are allocated to a treatment or a placebo arm by chance.Therefore, observed differences between the groups result solely due to the treatment. Yet, RCTs are time consuming, resource-intensive, and at times impossible to conduct due to ethical considerations (Lawlor et al., 2008).

To address these shortcomings, Mendelian Randomization (MR) was developed. MR is a causal inference method that uses genetic variants as instrumental variables (IVs) to investigate the causal effect of an exposure on an outcome. As genetic variants are inherited randomly at birth, Mendelian Randomization is considered a natural genetic counterpart to randomized controlled trials. Like RCTs, MR is less prone to reverse causation and confounding than observational studies. In contrast to RCTs, MR requires no intervention and can be performed even on cross-sectional epidemiological data, allowing for a more diverse study sample than the typically restricted populations in RCTs (Lawlor et al., 2008). Furthermore, MR is cost- and time-efficient, in contrast to RCTs which require longitudinal data to investigate causal effects, which becomes particularly challenging to estimate life-time causal effects (e.g., effect of diet on a long-term outcome such as cancer) (Lawlor et al., 2008). Yet, it is important to note that MR estimates are prone to other sources of bias. To yield accurate causal estimates, MR relies on three main assumptions (*Figure 1a*): First, the relevance assumption: the genetic variants are robustly associated with the outcome. Second, the exchangeability assumption: the genetic variants are not associated with any confounder of the exposure-outcome relationship. Third, exclusion restriction: the genetic variants are only associated with the outcome through the exposure, i.e., the relationship between the genetic variants and the outcome is fully mediated through the exposure (Lawlor et al., 2008) (*Figure 1a*). Whilst these are the standard assumptions, MR has other implicit assumptions, such as gene-environment equivalence (a change in the exposure should have the same effect on the outcome, regardless of whether it results from genetic or environmental variation (Sanderson et al., 2022)), causal effect linearity (the causal effect of an exposure on an outcome is the same for all levels of the exposure), and effect homogeneity (the obtained causal effect is the same for everyone (Sanderson et al., 2022)).

**Figure 1.**
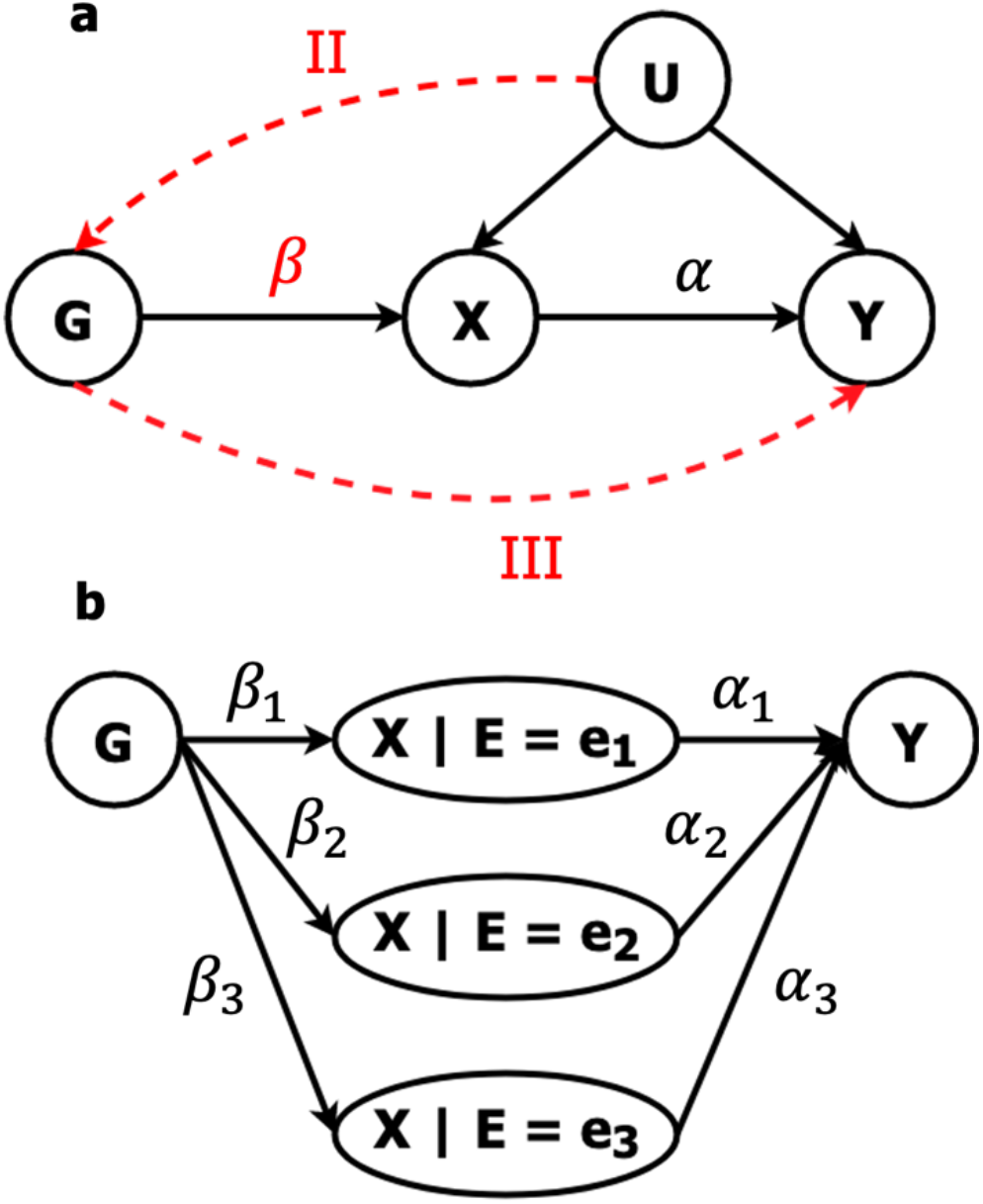
a: MR assumptions. G = genetic instruments, X = Exposure, Y = Outcome, U = Confounding variable ß = relevance assumption, II = exchangeability assumption, III = exclusion restriction. b: Interaction analysis as performed by Richardson and colleagues. By selecting genetic variants whose association with X depends on the interaction parameter E, different levels of X depending on E can be obtained. Lifetime Y is regressed on those levels of X in a uni- and multivariable fashion to investigate the causal effect of X on Y depending on the level of E.

In this work, we aim to address one plausible cause for the violation of the effect homogeneity assumption, by considering how environmental variables may modulate causal effects (even in presence of non-linear exposure-outcome relationships).

### Interactions in MR

Recent studies indicate that environmental factors (E) such as age may modulate the observed causal estimates of an exposure (X) on an outcome (Y). For example, Richardson and colleagues (2020) investigated if the effect of body size on different health outcomes is age dependent. After obtaining distinct genetic instruments for adult and childhood body size, they fitted both uni- and multivariable MR models on a range of outcomes. If the effect of early life body size is significant in the univariable MR framework, but not in the multivariable MR framework, it indicates that the association is likely mediated through adult body size (and vice versa). Whilst the effect of early life body size on coronary heart disease was fully mediated through adult body size, they observed a strong protective effect of larger childhood body size on breast cancer risk, independent of adult body size. Richardson’s approach allows for investigating the effect of the same exposure (e.g., body size) at different levels of the environment (e.g., age) on an outcome Y (e.g., coronary heart disease) (*Figure 1b*).

For Richardson’s approach, it is necessary to obtain IVs to separately instrument the exposure at different levels of the environment. This requirement implies two major challenges. First, the levels of the environment are defined arbitrarily. Richardson and colleagues considered childhood body size and adult body size because data was available for these two time points in the UK Biobank (UKBB). Depending on the available data, artificial stratification of the environmental variable E is necessary, which likely would impact the results. Second, it is necessary to obtain IVs that differ in their association with the exposure depending on the environment E. For the ranges of age, socioeconomic status, smoking, air pollution, TV time, and physical activity that are available in the UKBB, we only found evidence for at least one genome-wide significant gene-environment interaction effect for 22 out of 228 environment – exposure pairs. Therefore, the applicability of this approach is very limited.

### Interactions in presence of non-linear causal effects

It has long been known that interactions can arise from unspecified nonlinear effects of correlated variables (Cortina, 1993; Matuschek *S* Kliegl, 2018). In simpler terms, if the exposure term X and the effect modifying variable E are correlated and X^2^ is associated with the outcome Y, but the quadratic effect is not included in the regression model, the *X* · *E* interaction term may be overestimated. Whilst the reverse is also true (i.e., in presence of a true interaction, if only the quadratic effect is included in the model, but not the interaction, the quadratic effect may be overestimated), the present work focuses on obtaining accurate interaction estimates and only considers nonlinear effect estimates as a nuisance parameter. Considering non-linear effects is of relevance as they play a role in a wide range of causal exposure-outcome relationships. A common example is the J-shaped association between BMI and all-cause mortality (Berrington de Gonzalez et al., 2010), which has not only been reported in observational, but also in MR studies (Sun et al., 2019). Furthermore, Sulc and colleagues (2021) estimated the causal effects of four anthropometric traits (such as body fat percentage) on seven health biomarkers (such as systolic blood pressure) and found significant evidence for nonlinearity for most of the investigated causal effects (84%).

Thus, to address the abovementioned limitations, we developed 2SLS-I, an approach to estimate exposure interactions with a non-instrumentable environmental variable E (e.g., age, socioeconomic status, physical activity), while accounting for potential nonlinear causal effects. We suggest investigating interactions by instrumenting *X* · *E* with *G* · *E*, where G is a genetic instrument for the exposure X. The advantage of this approach is that it does not require the existence of genetic instruments for X that have different effects on X depending on the value of E and it allows for accounting for non-linear effects. In extensive simulation settings, we were able to obtain accurate main- and interaction estimates even in presence of sources of bias, such as linear and quadratic confounding, a quadratic exposure-outcome relationship, and a causal effect of the environment on the exposure. Application to a wide range of exposure (e.g. body fat percentage, education, and Vitamin D), environment (e.g. age, socioeconomic deprivation, physical inactivity), and outcome (e.g. fluid intelligence score, forced expiratory volume within 1 s, CRP) combinations in the UK Biobank revealed multiple relevant settings where the environmental variable significantly modulated causal effects. For example, we found that multiple causal effects are attenuated with older age, whilst socioeconomic deprivation and smoking, in tendency, exacerbated detrimental causal effects.

## Methods

All simulations and analyses were performed using R version 4.2.1 “Funny looking kid “ (R Core Team, 2022). For the application of 2SLS-I, Snakemake version 7.25.3 (Mölder et al., 2021) was used as a workflow manager.

### Simulations

All simulations were performed with a sample size of 10’000 and 500 repetitions. We simulated the data as follows.

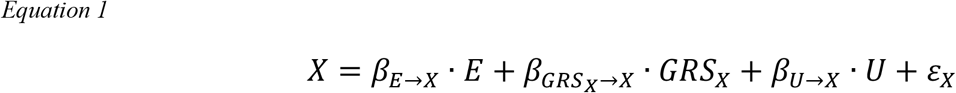

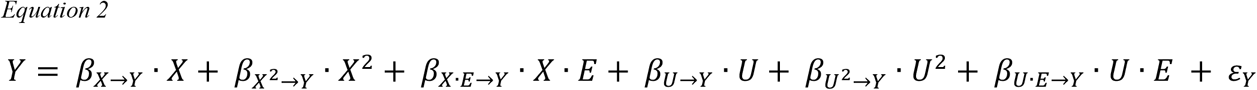

Where both E and U were drawn separately from a standard normal distribution. Thereby, E represents the non-instrumentable effect modifying environment (e.g. age), whilst U represents an unmeasured confounder. The GRS, representing a polygenic score for *X*, was drawn from a normal distribution with mean (*μ*) = 0 and variance 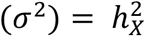, i.e., the variance of the GRS was set to be equal to the proportion of phenotypic variance explained by the polygenic score, which by default was set to 10%. The residual errors *ε*_*x*_ and *ε*_*Y*_ were drawn from a normal distribution with mean 0 and their respective variances were set to ensure that the overall variance of X and Y is equal to 1. For an overview of the varied variables, see *Table 1* and *Figure 2*.

**Table 1.**
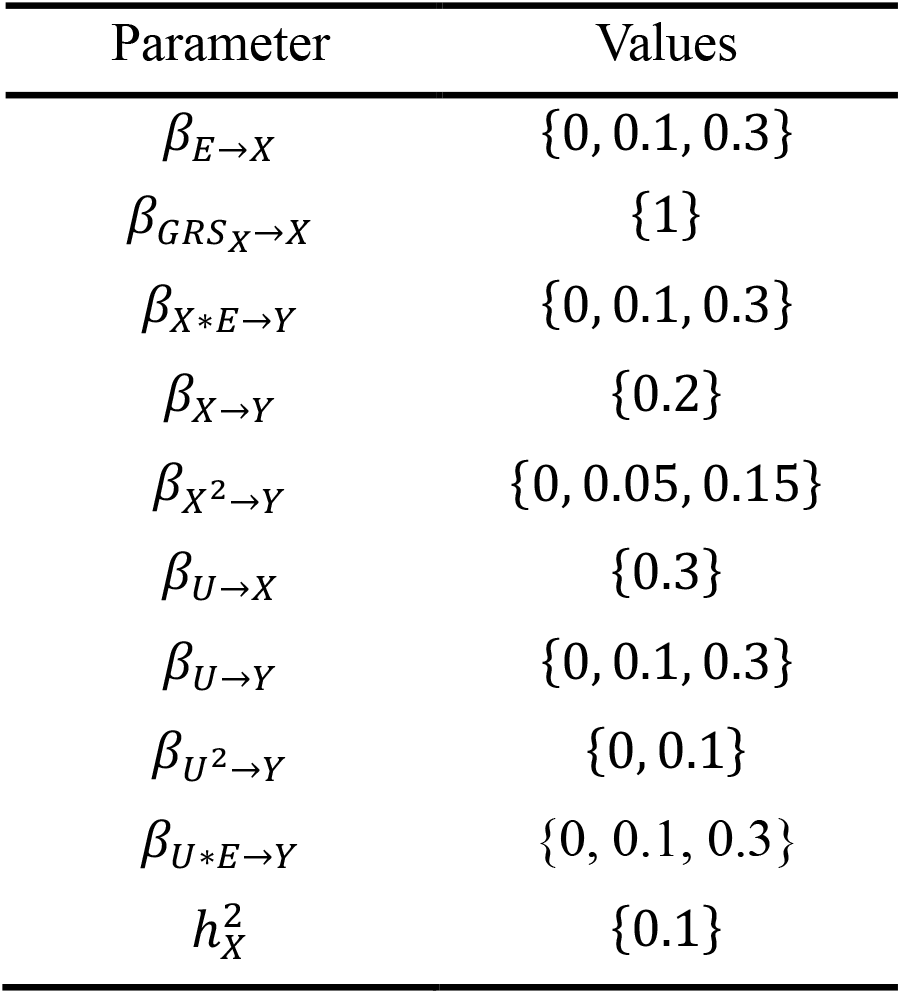
Overview of the varied and constant parameters in simulations.

**Figure 2.**
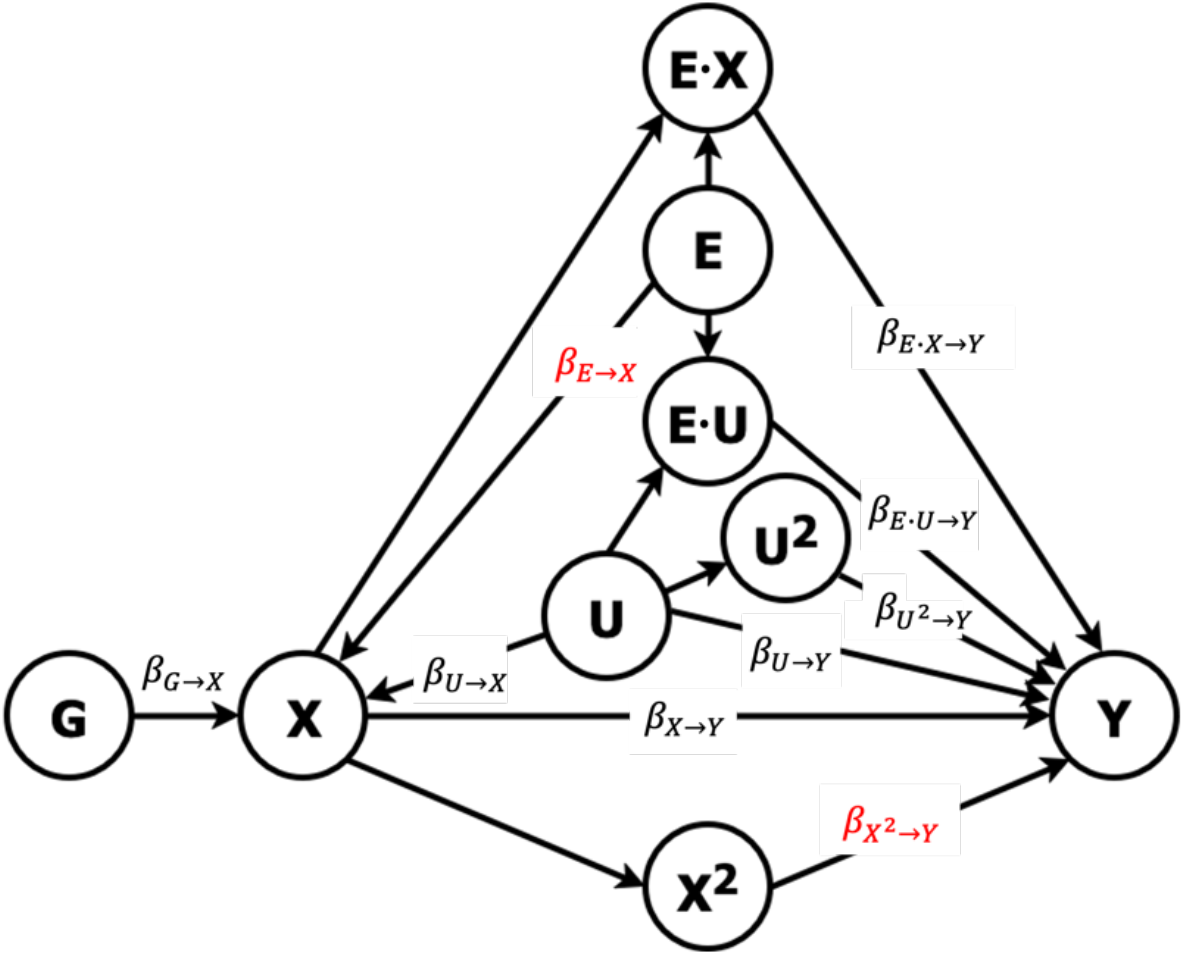
Data simulation settings. G = genetic instruments for exposure X, X = exposure, U = confounding variable, E = non-instrumentable environment, Y = outcome.

**Figure 3.**
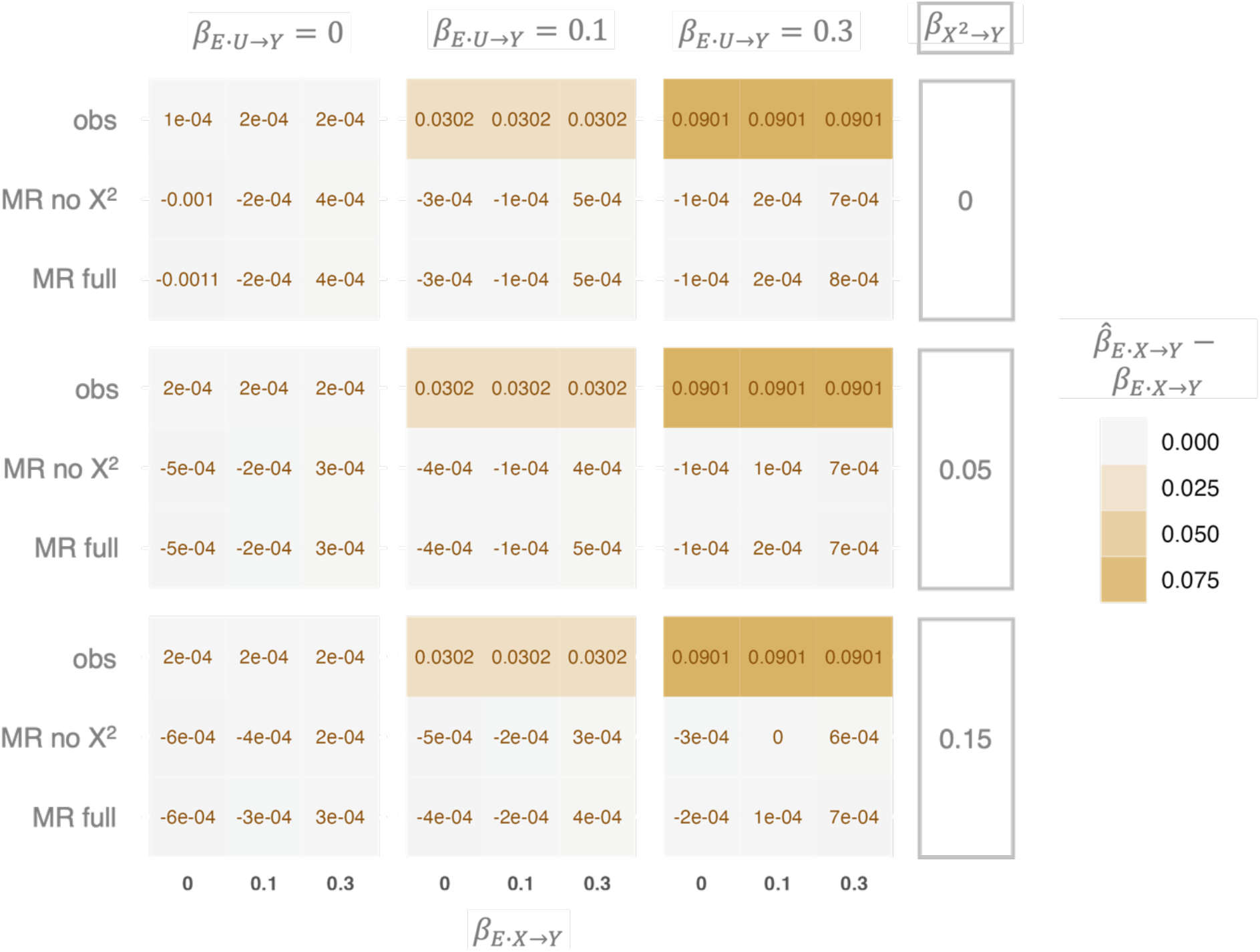
Deviation from the true simulated interaction effect for interaction estimates obtained from observational (obs) and two different MR models (MR no X^2^: no quadratic effect of X on Y fitted, MR full: full MR model) in presence of an interaction between the environment E and the confounder U and a quadratic effect of X on Y. The tile color indicates deviation of the interaction estimate from the simulated interaction, with darker colors indicating stronger deviations.

For all MR models, *β*_*U*→*Y*_ was kept at 0.3 and 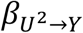 at 0.1, invoking linear and quadratic confounding. Four models were fitted to investigate the accuracy of the interaction estimate in different settings. An observational model, and three different MR models, where we once omitted the quadratic exposure term, once omitted the interaction term, and once fitted the full model (*Table 2*). In the main text, we present estimates from the full MR model.

**Table 2.** Models fitted in simulations. Y refers to the outcome variable, X to the observational exposure variable, E to the (non-instrumentable) environmental variable and 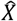 to the exposure variable when the effect is obtained using the GRS for X.

| Model | Formula |  |
| --- | --- | --- |
| Observational | $Y \sim X + E + X^2 + E^2 + X \cdot E$ | 207 |
| MR without interaction | $Y \sim \hat{X} + E + E^2 + \hat{X}^2$ | 208 |
| MR without quadratic | $Y \sim \hat{X} + E + E^2 + \hat{X} \cdot E$ | 209 |
| Full MR model | $Y \sim \hat{X} + E + E^2 + \hat{X}^2 + GRS \cdot E$ | 210 |

For the MR models, we instrument *X* · *E* with *G* · *E*, where G represents a genetic score based on the genome wide significant independent genetic instruments for X. Evidently, the MR assumptions need to be extended to *G* · *E* for it to be a valid instrument of *X* · *E*.

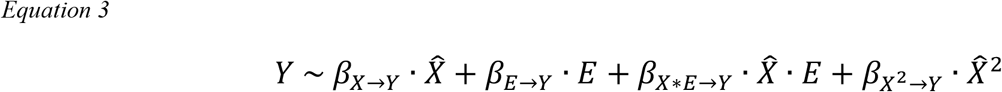

with

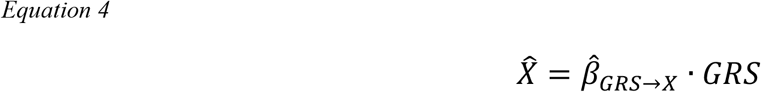

Model Formula

### Correction approach: 2SLS-I-corr

In simulations, we found a systematic bias of the interaction estimate if the causal effect modifying variable E had an effect on the exposure X and X^2^ had an effect on the outcome Y. From Equation 1 we obtain Equation 5.

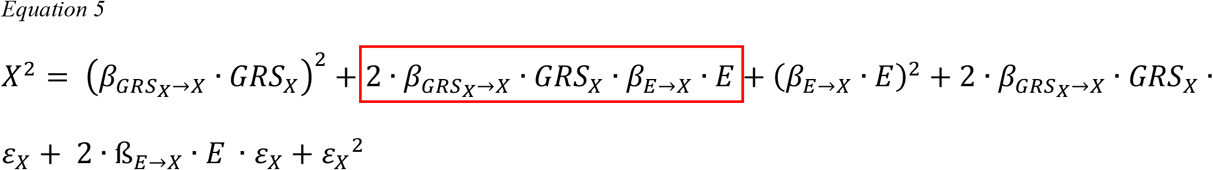

In Equation 5, it becomes evident that the MR interaction term *GRS* · *E* is also represented in *X*^2^, if *β*_*E*→*X*_ ≠ 0. As a consequence, the raw *GRS* · *E* estimate is biased if X is a function of E and X^2^ influences Y. From this, one can derive the following correction term.

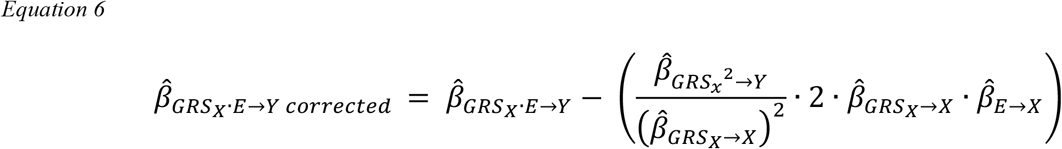

Where the term in parentheses represents the extent to which 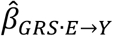 deviates from the true *β*_*GRS*·*E*→*Y*_ if 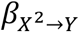 and *β*_*E*→*X*_ are not equal to 0. 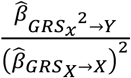 translates to the causal estimate of 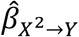. The remaining terms represent the extent to which the true interaction is overestimated in response to the quadratic effect of X on Y (i.e. the extent to which *X*^)^ is a function of *GRS* · *E*), as visible in Equation 5 (marked in red). The full correction term from Equation 6 can be simplified as follows.

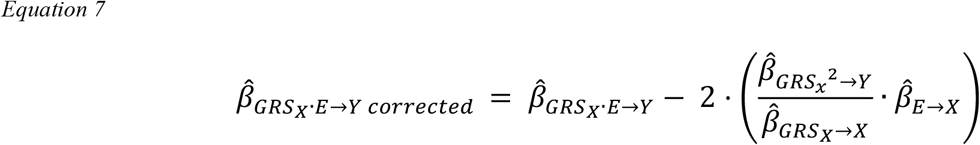

From Equation 7, using the first order Taylor expansion-based approximation of the variance of ratios, we can derive the variance of the corrected interaction term as follows.

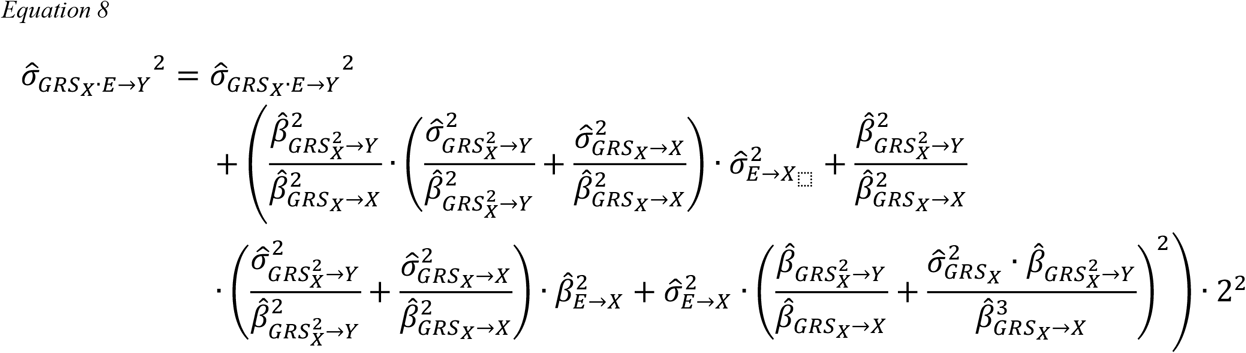

By setting 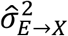 to 0 (i.e. ignoring the variance in the E-to-X effect estimation), Equation 8 can be approximated as follows.

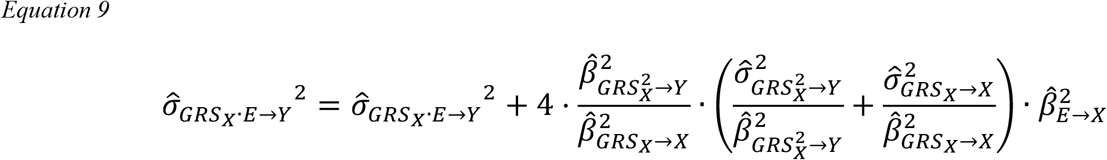

### Power analysis

The accuracy and power of the corrected interaction estimate was assessed. For the power analysis, *β*_*E*→*X*_ was set to 0.2, whilst the amount of variance that the GRS explained in 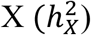, the true interaction (*β*_*GRS*·*E*→*Y*_), the quadratic effect of X on 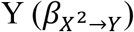, and the sample size were varied systematically. The settings for the power analysis are listed in *Table 3*.

**Table 3.**
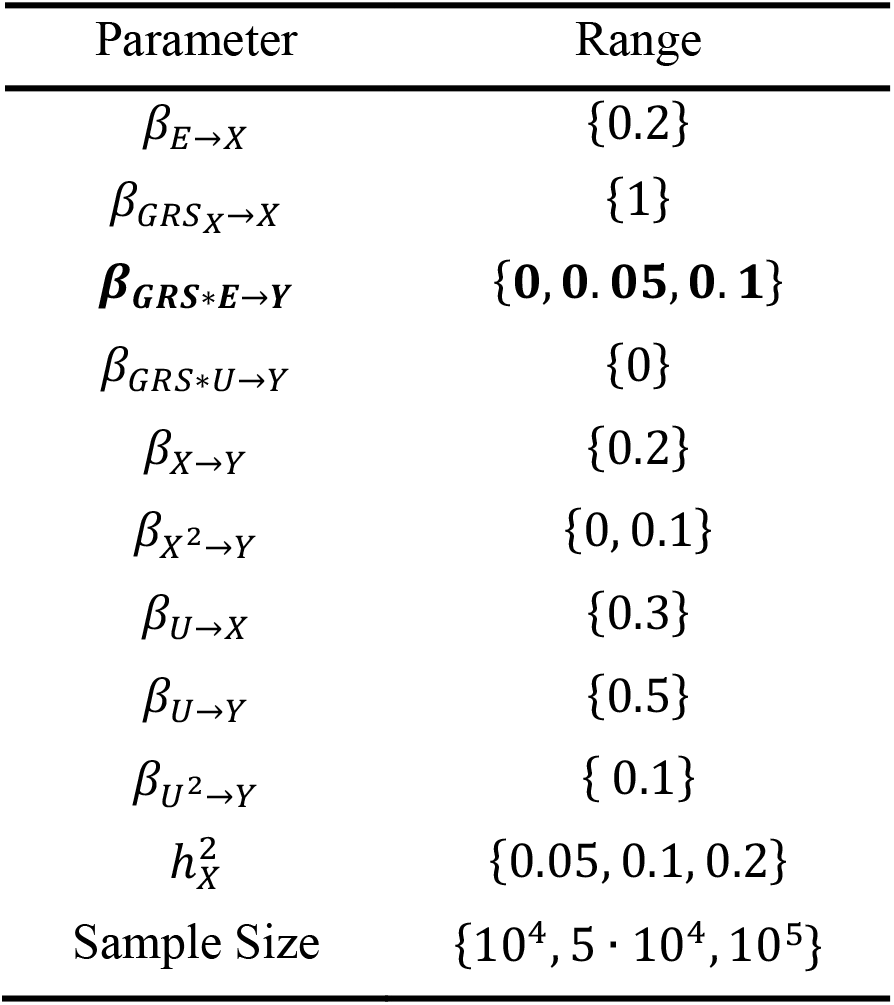
Simulation settings of the statistical power analysis.

### Application to UK biobank

We applied 2SLS-I to the UK biobank (UKBB) to investigate if interactions with environmental variables occur and how prone they are to bias induced by nonlinear effects of the exposure on the outcome.

### Cohort description

UKBB is a volunteer-based biomedical cohort of ∼500’000 individuals from the general UK population (Sudlow et al., 2015). The data was accessed through application number 16389. We selected unrelated, white British participants, for whom the inferred and reported sex aligned, leading to an initial sample size of n = 3370’392.

### Phenotype selection

We selected environmental variables which are difficult to instrument genetically but are likely to modulate causal relationships, namely age, current smoking, physical inactivity (obtained using accelerometer data, sedentary behaviour, “PA: sed ”), air pollution (Nitrogen dioxide (“AP: NO_2_ ”), time spent watching TV (“TV time ”), and socioeconomic deprivation (Townsend deprivation index, “TDI ”)). A range of outcome traits was selected to address different health-, and cognitive variables, namely: fluid intelligence score (FIS), reaction time (RT), systolic blood pressure (SBP), low-density lipoprotein cholesterol (LDL), hand grip strength (HGS), forced expiratory volume within 1 second (FEV1), and C-reactive protein (CRP). For each of the outcome traits, we searched the literature and the EpiGraphDB (Liu et al., 2021) to obtain exposures that might have a causal effect on any outcome of interest. Exposure traits were considered for inclusion if they were strictly numeric, conceptually different from the outcome of interest (e.g. we removed HDL cholesterol as exposure for LDL cholesterol as the two variables are tightly related) and had at least one genome wide significant variant. If a numeric alternative for a categorical trait was available, the numeric alternative was considered (e.g. instead of *hypertension*, we used *systolic blood pressure*). If multiple potential exposures covered conceptually very similar traits, only one was selected. For an overview of the selected exposure-, effect modifying environment-, and outcome traits, and the according UKBB field IDs, see *Supplementary Tables 1-3*. Exposure and outcome phenotypes were corrected for age, age^2^, sex, sex*age, and 40 genetic PCs and exposure phenotypes were inverse rank normal transformed (IRNT) (McCaw et al., 2020). Potentially effect modifying environmental phenotypes were corrected for age, age^2^, sex, and age*sex, except age (only corrected for sex). Effect modifying environment and outcome phenotypes were standardized to have zero mean and unit variance.

### SNP selection and GRS calculation

For each exposure phenotype, we accessed summary statistics (Neale, 2017) and filtered for SNPs with a p-value < 10^−4^. Next, we clumped the selected SNPs using PLINK version 1.9 to obtain independent genetic variants for MR. For the remaining variants, we reassessed their association with the IRNT exposure phenotype of interest whilst correcting for age, sex, age^2^, and 40 PCs. For the GRS, we only kept SNPs which were genome wide significantly associated (p < 5*10^−8^) with the exposure in our sample. The selected SNPs were used to obtain the GRS for each exposure of interest. The GRS was calculated as follows: 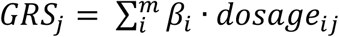, where *m* is the number of genome wide significant SNPs, *β*_*i*_ is the obtained effect size of *SNP*_*i*_ on the exposure of interest and the *dosage*_*ij*_ is the number of copies of the coded allele for SNP *i* in individual *j* (Collister et al., 2022). The obtained GRS was scaled to have zero mean and unit variance. We further assessed if there was evidence for an interaction between the GRS and the environment E on the exposure X (level 1 interaction) at p < 0.05. We observed this in 77 out of 228 (33.8%) of the settings (of which 38 survive Bonferroni correction). In-depth analysis of the role of level 1 interactions revealed a negligible role for level 1 interactions (for details on simulations and application, see Supplementary Material *Level 1 interactions*).

### Inclusion of extended GRS (GRS_ext_) to boost statistical power

As interaction analyses require strong effects and a large sample size to obtain sufficient power, and the application of the quadratic effect correction further reduces the power to detect true interactions, we additionally performed MR with an extended GRS (GRS_ext_) to boost for power. GRS_ext_ was calculated for all selected exposure phenotypes as described in Privé et al. (2022). All analyses were performed as for the strict GRS.

### Mendelian Randomization Models

Next, the GRS was used to instrument the effect of each exposure on each outcome whilst considering the effect modifying environmental variables of interest. We fitted four different models for each setting, including the full model: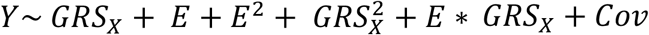, a model without the quadratic term: *Y* ∼ *GRS*_*X*_ + *E* + *E*^2^ + *E* · *GRS*_*X*_ + *Cov*, a model without the interaction term: 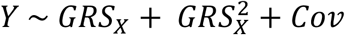, and a minimal model: *Y* ∼ *GRS*_*X*_ + *Cov*, where Y denotes the outcome of interest, the GRS is the previously obtained GRS for X, E is the effect modifying environmental variable of interest, and Cov translates to the covariates in the model. For all settings where E was not age, the covariates were age, age^2^, sex, and relevant medication. If the environmental variable was age, the age-related terms were excluded from the covariates. Only relevant medications were corrected for, obtained using UK Biobank data fields 6153 and 6177 for women and men, respectively. For an overview of the applied medication correction, see *Supplementary Table 1*. We used the ratio estimates for the causal effects (X-to-Y and X×E-to-Y), i.e. 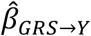 and 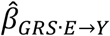 were divided by 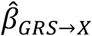, whilst 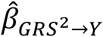 was divided by 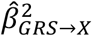.

### Correction of the interaction

For the correction of the interaction (2SLS-I-corr), separate models were fitted to obtain 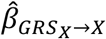 and 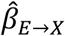 (and the according standard errors to obtain the variance). X was inverse-normal rank transformed and E was z-standardized, and the models were corrected for the same covariates as the previously discussed MR models (depending on the exposure X and the environment E: age, age^2^, sex, medication). 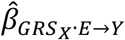 and 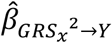 were obtained from the full MR model described above. 2SLS-I-corr was only applied as discussed in the section *Correction approach: 2SLS-I-corr* if 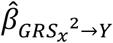 was nominally significant (p < 0.05), otherwise the correction would just reduce power due to the increased estimator variance.

### Interaction tier score

To gauge the reliability of the obtained interactions using both, the strict GRS and GRS_ext_, we developed a rating strategy. For each interaction, we assessed whether 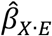 differed significantly from 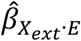 From this, tiers were computed as follows.

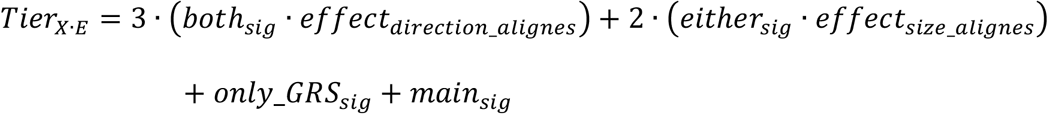

Where all variables are binary. *both*_*sig*_ is equal to 1 if both, the GRS and the GRS_ext_ yield significant (at p < 0.001) interaction estimates, and 0 otherwise. *effect*_*direction_alignes*_ is 1 if the sign of the effects obtained by the GRS and the GRS_ext_ agrees, and 0 otherwise.

*effect*_*size_ alignes*_ is 1 if the effect sizes obtained by the GRS and the GRS_ext_ do not differ significantly (p > 0.05), and 0 otherwise. *only_GRS*_*sig*_ is 1 if the GRS yielded significance at p < 0.001, but the GRS_ext_ did not, and the effect size obtained by the GRS does not align with the effect size obtained by the GRS_ext_. *main*_*sig*_ is 1 if there is Bonferroni corrected significant evidence for a causal effect of the exposure on the outcome. This approach gives the highest confidence for interactions which yield significance for both, the GRS and the GRS_ext_, and if on top of that the sign of the obtained interaction effect aligns. If either the GRS or the GRS_ext_ yields a significant effect (at p < 0.001), and the obtained effects do not differ significantly (p > 0.05), the interaction would be classified as tier 2. If only the GRS yields a significant interaction (at p < 0.001), but the GRS_ext_ estimate does not agree, the interaction is classified as tier 1. By this we aim to account for the higher reliability of the GRS in comparison to the GRS_ext_. Finally, if an interaction yielded a tier score > 0, we increased the tier score by +1 if we found significant evidence (p_bonferroni_ < 0.05) for a main effect of the exposure on the outcome.

### Sensitivity analyses

To ensure the robustness of our findings, we performed extensive sensitivity analyses validating our results.

For example, we assessed whether there was evidence for a stronger effect of the exposure on the environment than vice versa and flagged up such interactions as this circumstance may lead to a (collider) bias in the interaction estimate. The according interactions are shaded out in the respective results plot. For details of the analysis, see Supplementary Material, *Exposure on environment effects*.

Furthermore, we assessed the robustness of our results to inverse rank normal transforming the outcome phenotypes. For details of the analysis, see Supplementary Material, *Inverse rank normal transformed outcome phenotypes*.

Finally, to investigate if some interactions are only picked up due to method specific features of 2SLS-I, we assessed the same settings in an extended doubly ranked stratification (Tian et al., 2023) framework (DRS). Whilst the doubly ranked stratification method was developed to investigate non-linear exposure-outcome relationships, it can be adapted to allow for investigating interactions by stratifying by an environmental variable (instead of the exposure value) adjusted by the exposure’s instrument and regressing the obtained MR estimates on the predefined strata midpoint value. For a detailed effect comparison between the two methods, see Supplementary Material, *Replication analysis*.

## Results

### Simulations

#### Biased interaction effects for the uninstrumented regression model

Before assessing the accuracy of interaction effects in a Mendelian Randomization (MR) framework, we investigated under which circumstances MR may yield an advantage over observational models. We found that interactions (in contrast to main effects 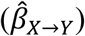 and quadratic effects 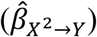, respectively) are robust against linear (*β*_*U*→*X*_ = 0.3, *β*_*U*→*Y*_ = 0.3) and quadratic 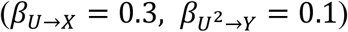 confounding (M_diff_ = -0.00007, SD_diff_ = 0.009). Yet, the observational model yielded biased interaction estimates in presence of an interaction between the confounder (U) and the environment (E) (*β*_*U*→*X*_ = 0.3, *β*_*U*·*E*→*Y*_ = 0.3, M_diff_ = 0.9, SD_diff_ = 0.0099). In contrast, the interaction estimates obtained from MR were accurate even in the strongest settings of confounding (*β*_*U*→*X*_ = 0.3, *β*_*U*→*Y*_ = 0.3, *β*_*U*·*E*→*Y*_ = 0.3) (M_diff_ = -0.0003, SD_diff_ = 0.036).

#### Biased interaction estimates in presence of quadratic effect of X on Y and effect of E on X

We assessed the accuracy of the interaction estimates in an MR framework using simulations. Data was simulated with a constant effect of the exposure on the outcome (*β*_*X*→*Y*_ = 0.2), whilst the levels of the true interaction between the environment and the exposure (*β*_*E*·*X*→*Y*_), the quadratic effect of the exposure on the outcome 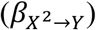, and the effect of the environment on the exposure (*β*_*E*→*X*_) were varied systematically (for simulation settings, see *Table 1*).

We observed that if both the association between the environmental variable E and the exposure X (*β*_*E*→*X*_) and the quadratic effect of the exposure X on 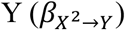 deviated from 0, the obtained raw interactions (2SLS-I-raw) between GRS and E on 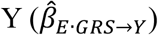,were systematically biased. For the most extreme simulation settings (*β*_*E*→*X*_ = 0.3 and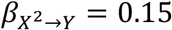), 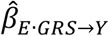was overestimated by 0.089 (SD = 0.035) on average.

In absence of a quadratic effect of X on 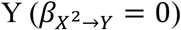,even a strong association between the environmental variable E and the exposure X (*β*_*E*→*X*_ = 0.3) did not lead to any bias using 2SLS-I-raw (M_diff_ = -0.0009, SD_diff_ = 0.033). Vice versa, if *β*_*E*→*X*_ was set to 0, the obtained raw interaction estimates did not deviate from the true interaction, even in presence of a very strong quadratic effect of the exposure X on 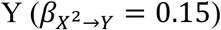 (M_diff_ = -0.0009, SD_diff_ = 0.034) (*Figure* 4).

**Figure 4.**
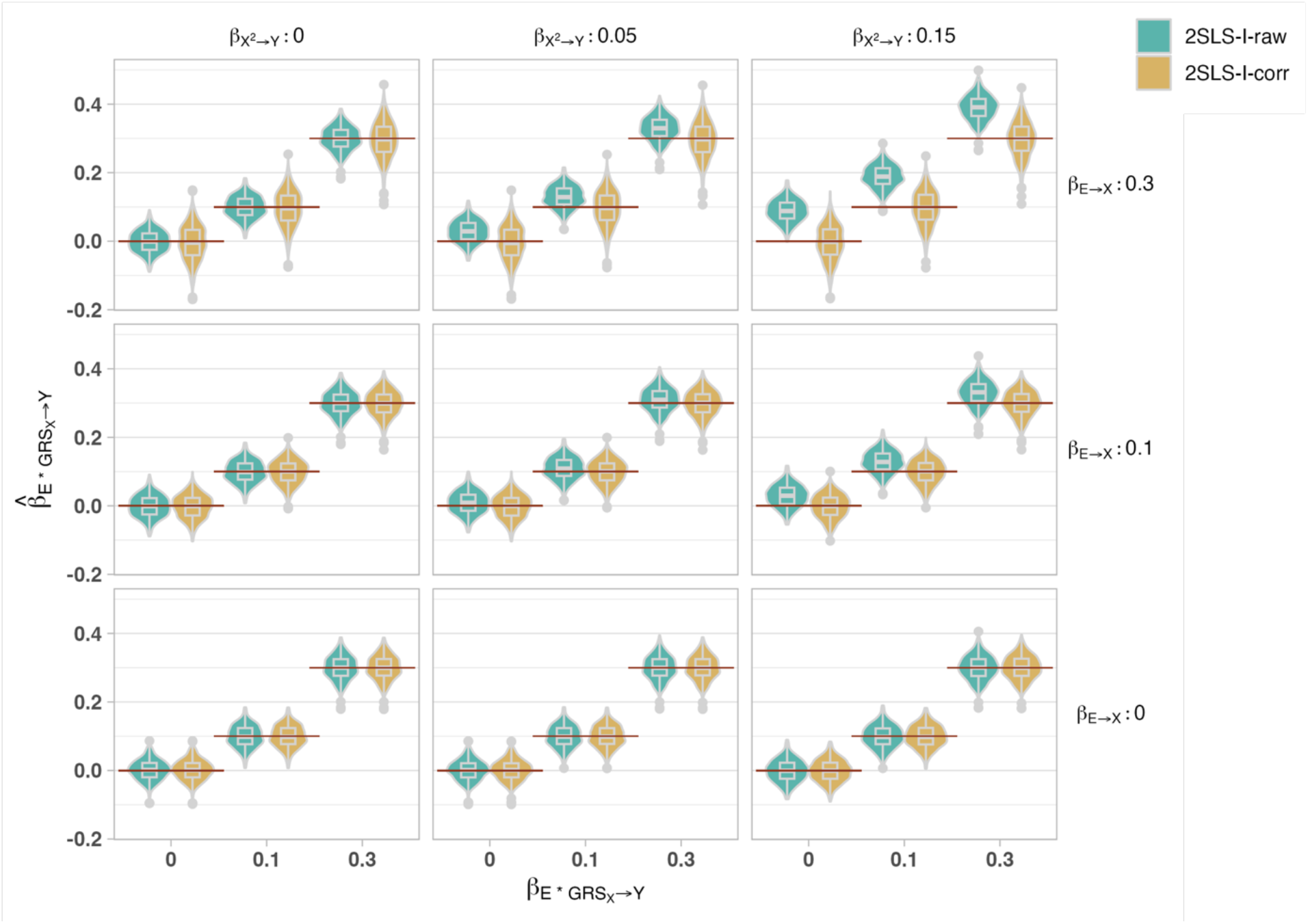
The obtained raw (2SLS-I-raw, blue) and corrected (2SLS-I-corr, yellow) interaction estimates for different levels of 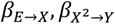and 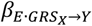The red lines indicate the different levels of the simulated interaction effect 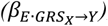,also indicated the x-axis. We observed accurate raw interaction estimates as long as either the environment-exposure or the exposure outcome quadratic effect is absent (β_E→X_ = 0 or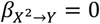) (corresponding to plots in the bottom row and left column). Deviation of bothβ_E→X_ and 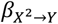 from 0 led to an overestimation of the raw interaction (in blue, 2SLS-I-raw) between GRS and E on Y (top right). This bias was successfully attenuated in the corrected interaction estimates (in yellow, 2SLS-I-corr). The elements of the boxplot are as follows: center line: median, box limits (lower and upper hinges): quartiles (25^th^ and 75^th^ percentile), upper whisker: largest value no further than 1.5 times the interquartile range (IQR) from the hinge, lower whisker: smallest value no further than 1.5 times the IQR from the hinge. Data beyond the end of the whiskers are plotted individually as dots.

**Figure 5.**
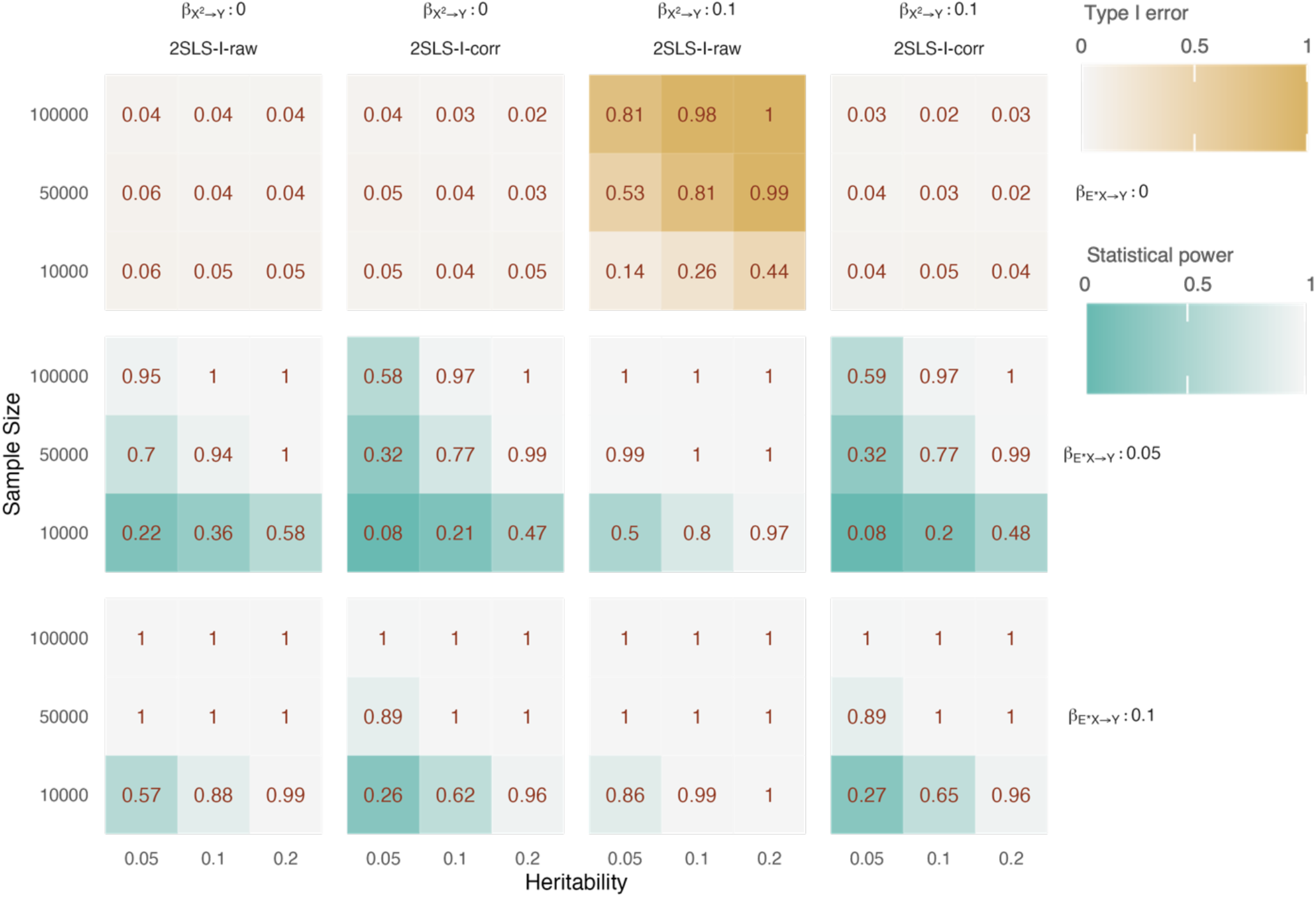
Analysis of the type I error (T1E) and power of raw (2SLS-I-raw) and corrected (2SLS-I-corr) interaction estimates in absence and presence of bias. The first row illustrates the type I error, as the true interaction between E and X (β_E·X→Y_) was set to 0. In absence of any source of bias (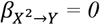, first and second column), the T1E of both, 2SLS-I-raw and 2SLS-I-corr are close to 5R. In presence of a quadratic effect of X on Y (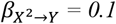, third and fourth column), the raw interaction estimates are nominally significant in up to 100R of the cases in absence of a true interaction. This effect is successfully attenuated in the corrected interaction estimates, where the T1E does not exceed 5R. In presence of a weak interaction (middle row), 2SLS-I-corr showed reduced power in comparison to 2SLS-I-raw in absence of bias. C: Lastly, the power of 2SLS-I-corr reached a ceiling effect in presence of a strong interaction (β_E·X→Y_ = 0.1) with a sample size ≥ 50.000 and 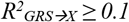 (bottom column).

### Accuracy of 2SLS-I-corr

As described in the section *Correction approach: 2SLS-I-corr*, we investigated the source of bias and developed a method to correct for the bias observed if both the association between the environmental variable E and the exposure X (*β*_*E*→*X*_) and the quadratic effect of the exposure X on 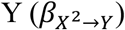 deviated from 0 (2SLS-I-corr). 2SLS-I-corr yielded unbiased interaction estimates independent of the simulated settings. Most importantly, 2SLS-I-corr allowed for accurate interaction estimates even in presence of both a strong association between E and X (*β*_*E*→*X*_ = 0.3) and a strong quadratic effect of X on 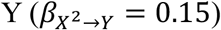 (M_diff_ = -0.003, SD_diff_ = 0.05). As expected, when both the quadratic effect of X on 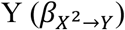,and the association between the effect modifying environment, E, and the exposure X (*β*_*E*→*X*_) were set to 0, 2SLS-I-corr yielded accurate estimates of the interaction (M_diff_ = 0.0001, SD_diff_ = 0.033). In presence of a strong quadratic effect of X on 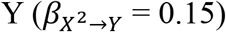, and in absence of an association between E and X (*β*_*E*→*X*_ = 0), the estimates from 2SLS-I-corr did not deviate from the simulated interaction (M_diff_ = -0.0008, SD_diff_ = 0.034). It is worth noting that we observed an increase in the standard error (SE) for 2SLS-I-corr in comparison to the SE of the raw interactions (2SLS-I-raw) (M = 0.0081, SD = 0.0094, p < 0.0001, 95% CI [0.0080, 0.0083]). We fitted a multiple linear regression to predict the difference in the SE between 2SLS-I-corr and 2SLS-I-raw based on 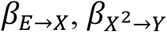, and *β*_*E*·*X*→*Y*_. This resulted in a significant model, F(3, 13496) = 87140, p < 0.0001, R^2^ = 0.95. The increased SE was positively associated with the strength of the association between the environmental variable, E, and the exposure X (*β*_*E*→*X*_, ß = 0.074, SE = 0.0001, p < 0.0001). Furthermore, the difference between the SE of the raw and corrected interaction was slightly reduced in presence of a stronger interaction between E and X (*β*_*E*·*X*→*Y*_, ß = -0.0004, SE = 0.0001, p = 0.0058). The strength of the quadratic effect of the exposure on the outcome 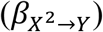 did not significantly contribute to the increase in variance observed in the corrected interaction estimates (ß = 0.00005, SE = 0.0003, p = 0.87).

To assess the accuracy of the estimator of the SE of 2SLS-I-corr, we compared our analytically derived SE with the empirical SE of the corrected interaction estimates. For comparison, we did the same for the raw interaction estimates, separately for each simulation setting. Across all settings, we found that the analytical SE agreed well with the empirical SE for both, the 2SLS-I-raw and 2SLS-I-corr. The raw model SE did not significantly differ from the raw empirical SE (mean difference = 0.002, and a similar observation was made for the corrected model SE (mean difference = 0.00098). This confirms that our analytical formula for the variance of the corrected effect is unbiased.

### Power analysis of 2SLS-I

To investigate the impact of our correction approach in hypothesis testing, we compared the type I error (T1E) and power of the 2SLS-I-corr with that of the 2SLS-I-raw. In these simulations, the association between the environmental parameter E and the exposure X (*β*_*E*→*X*_) was set to 0.2, whilst the true interaction (*β*_*E*·*X*→*Y*_), the quadratic effect of the exposure X on the outcome 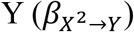,the amount of variance of X that is explained by the 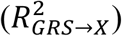, and the sample size were varied systematically (for the simulation settings, see Table 3).

In absence of a true interaction (*β*_*E*·*X*→*Y*_ = 0) and without the source of bias 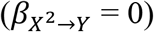, the (5% nominal level) T1E of 2SLS-I-corr (0.039 (SD = 0.01)) was comparable with the T1E of 2SLS-I-raw (0.046 (SD = 0.0089)). In presence of the source of bias 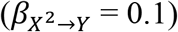, we observed a T1E of up to 1 for 2SLS-I-raw (on average 0.66 (SD = 0.33)). Importantly, even with a small sample size of 10.000 and a 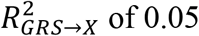 of 0.05, the T1E was 0.14 for the raw interaction in presence of bias. 2SLS-I-corr attenuated the T1E to a maximum of 0.05 (average of 0.035 across settings (SD = 0.0089)).

In presence of a weak interaction (*β*_*E*·*X*→*Y*_ = 0.05) and in absence of bias, 2SLS-I-raw had a power of more than 0.7 in all settings where the sample size was larger than 10.000 (M = 0.75, SD = 0.3). In contrast, 2SLS-I-corr only exceeded the power of 0.7 if both the sample size was larger than 10.000 and the amount of variance in X explained by the GRS exceeded 0.05 (M = 0.599, SD = 0.35). This reduction in power of the 2SLS-I-corr in comparison to the 2SLS-I-raw results from the increased variance in response to our additive correction in comparison to the raw interaction estimates.

In presence of a strong interaction, the power of 2SLS-I-raw exceeded 0.8 in all settings except if the 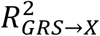 and the sample size were set to 0.05 and 10.000, respectively (M = 0.94, SD = 0.14). For 2SLS-I-corr, the power exceeded 0.8 in all settings except if the 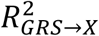 was smaller than 0.2 and the sample size was limited to 10.000 (M = 0.86, SD = 0.25).

In summary, 2SLS-I-corr has a much better controlled type 1 error than the 2SLS-I-raw whilst maintaining considerable power to detect true interactions if 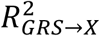 exceeded 0.05 and for sample size > 10.000 participants.

### Application

To elaborate the relevance of environmental moderators of causal effects, we investigated how a range of not genetically instrumentable environmental parameters (age, air pollution (NO_2_), sedentary behaviour, socioeconomic deprivation (Townsend Deprivation Index, TDI), smoking, and time spent watching TV (TV time)) modulate causal relationships between different health- and lifestyle-parameters. As simulations (see section *Biased interaction estimates in presence of quadratic effect of X on Y and effect of E on X*) indicated that non-linear effects of the exposure on the outcome may lead to spurious interaction effects, corrected interaction estimates (2SLS-I-corr) were considered in presence of a nominally significant quadratic effect of the exposure (215 out of 1274, 16.9%). For all other settings, raw interaction estimates (2SLS-I-raw) were considered, in order to maximise discovery power. To account for the limited power to detect true interactions, we replicated our analyses using an extended genetic risk score (GRS_ext_) in addition to the strict genetic risk score (where the GRS_ext_ is based on all SNPs that contribute to an exposure whilst the strict GRS is only based on independent SNPs that are genome wide significantly associated to an exposure). Evidence for interactions were classified into tiers from 0 to 4 to account for different levels of confidence (see section *Interaction tier score*), considering significance (of both, the GRS and the GRS_ext_), robustness of the interaction effect, and presence of a main effect.

Of the effect modifying environmental parameters, age, TDI, and smoking modulated the most causal relationships (n_age_ = 60 and n_TDI_ = 36, n_smoking_ = 22, respectively). In particular, we found a reduction in strength for a range of causal effects with increasing age. For example, the causal effect of height on hand grip strength (HGS) (ß = 0.22, SE = 0.0037, p < 10-^323^), was significantly attenuated with higher age (Tier score = 4, ß_GRS*E_ = -0.019, SE_GRS*E_ = 0.0037, pval_GRS*E_ < 10^−6^, *Figure 8a*).

For socioeconomic deprivation, we found that most interactions were associated with an exacerbation of the causal effects. For example, the negative effect of systolic blood pressure on forced expiratory volume within the first second (FEV1) (ß = -0.099, SE = 0.013, p < 10^−13^), was intensified for people exposed to higher levels of socioeconomic deprivation (Tier score = 3, ß_GRS*E_ = -0.021, SE_GRS*E_ = 0.013, p_GRS*E_ = 0.105, ß_GRSext*E_ = -0.022, SE_GRSext*E_ = 0.0041, p_GRSext*E_ < 10^−7^, *Figure 8b*).

Furthermore, we found evidence that smoking modulates 22 causal effects. For example, the positive causal effect of Gamma-glutamyl transferase (Gamma GLT) on CRP (ß = -0.107, SE = 0.006, p < 10^−76^) seems to be intensified in smokers (Tier score = 4, ß_GRS*E_ = 0.019, SE_GRS*E_ = 0.006, p_GRS*E_ < 0.001, ß_GRSext*E_ = 0.018, SE_GRSext*E_ = 0.004, p_GRSext*E_ < 10^−5^). Yet, it is worth noting that the interaction between smoking and Gamma GLT is scale dependent (i.e. the effect may be driven by individuals with very high CRP levels), as it yielded a tier score of 0 if the outcome was inverse-rank normal transformed.

Out of 19 causal effects that were modulated by air pollution (NO_2_), 8 affected the outcome CRP and 5 the outcome Forced Expiratory Volume (FEV1). For example, we observed that the effect of Education on FEV1 (ß = 0.208, SE = 0.016, p < 10^−38^) was significantly intensified for people who live in areas with higher air pollution (Tier score = 4, ß_GRS*E_ = 0.060, SE_GRS*E_ = 0.016, p_GRS*E =_ 0.0002, *Figure 8c*).

Sedentary behaviour, defined as time spent inactively, measured using an accelerometer device, mostly intensified causal effects on CRP (7 out of 8 interactions with a tier score > 0). For three of these interactions, we found evidence that physical inactivity may act as a collider between the exposures (BFP, education, and water mass) and the outcome (CRP), which likely leads to a bias in the interaction estimates. In addition, we observed that the effect of grip strength, cystatin C, Na in Urine, and SHB on CRP are significantly intensified in people who spend more time physically inactive. Finally, there was one setting where sedentary behaviour modified an effect on an outcome that is not CRP, namely the interaction with HbA1c on reaction time (tier = 1).

For time spent watching TV (TV time), we observed that the causal effect of Cystatin C on CRP (ß = 0.032, SE = 0.0061, p < 10^−7^) is intensified in people who spend more time watching TV (Tier score = 3, ß_GRS*E_ = 0.017, SE_GRS*E_ = 0.0061, p_GRS*E_ = 0.004, ß_GRSext*E_ = 0.02, SE_GRSext*E_ = 0.0034, p_GRSext*E_ < 10^−8^, *Figure 8d*). Noteworthily, the interaction between TV time and Cystatin C on CRP seems to be scale dependent (i.e. to some extent driven by individuals with very high levels of CRP), as the interaction on the IRN-transformed outcome phenotype yielded a tier score of 0.

For detailed results, see *Figure 6*, *Figure 7, Figure 8, Table 4*, and *Supplementary Table 4*.

**Table 4.**
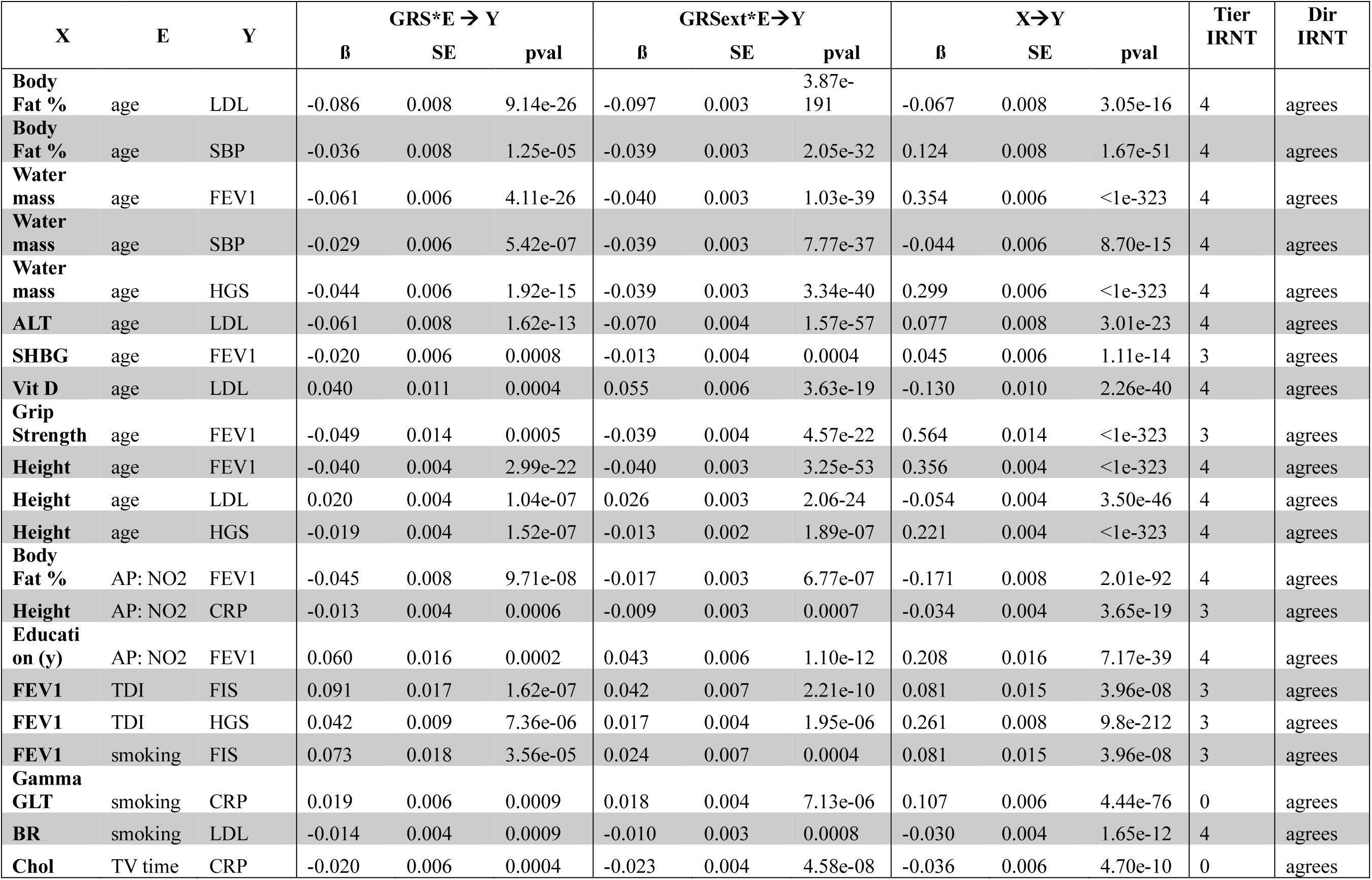
Overview of the tier 4 interaction effects with no evidence for a strong exposure-to-environment effect. Tier IRNT indicates the tier score obtained when the outcome was inverse rank normal transformed. Dir IRNT agrees in all settings, meaning that both, the strict and the extended GRS yielded an interaction effect pointing in the same direction as the interaction effect obtained for the z-standardized outcome, when the outcome was inverse rank normal transformed. Yet, there are two settings where the strength of the effect was potentially driven by individuals on the extreme end of the scale for CRP, as these interactions were attenuated to a tier score of 0 when interaction analysis was performed using the IRN-transformed outcome phenotype. AP: NO2: Air Pollution, Nitrogen Dioxide, CRP: C-reactive protein, FIS: Fluid intelligence score, HGS: Grip strength, FEV1: forced expiratory volume within one second, LDL: low density lipoprotein, SBP: systolic blood pressure, TDI = Townsend deprivation index.

**Figure 6.**
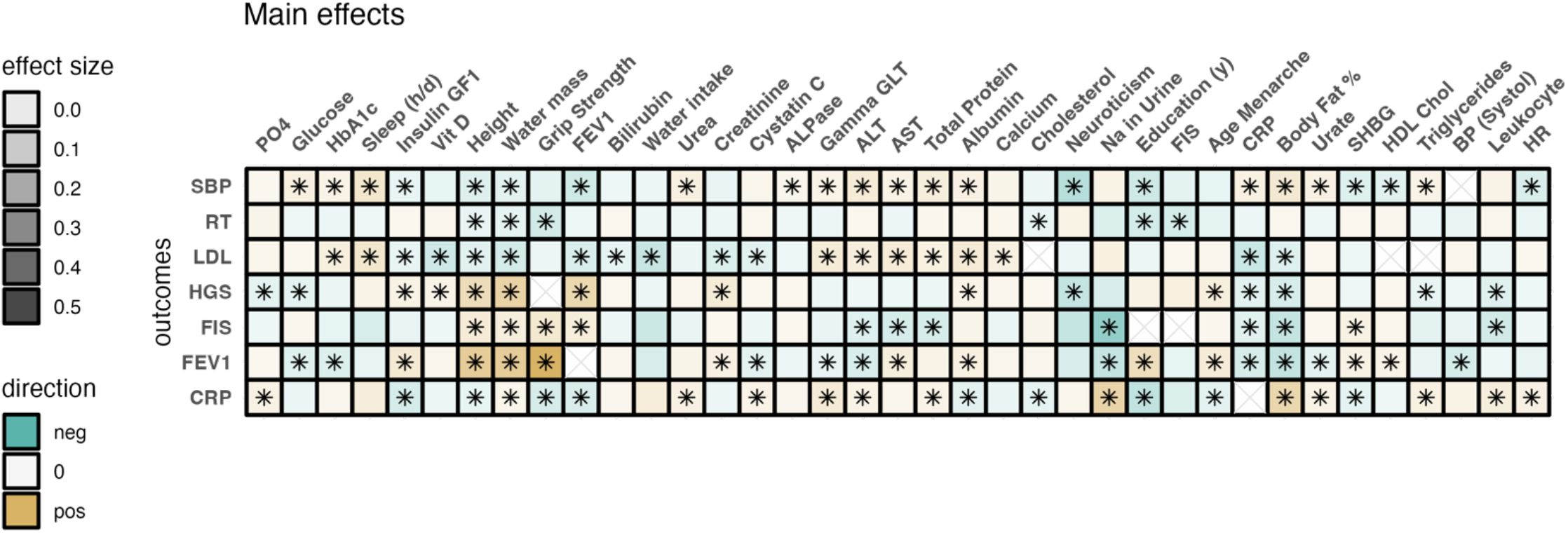
Causal effects of a range of exposures (x-axis) on a range of outcomes (y-axis), obtained from the minimal MR model. The color of the tiles indicates the direction of the main effect, whilst the opacity indicates the strength of the main effect. Bonferroni corrected significant effects are marked with a star. ALT: Alanine Aminotransferase, AST: Aspartate Aminotransferase, BP: Blood pressure, CRP: C-reactive Protein, FEV1: Forced Expiratory Volume within 1 second, FIS: Fluid Intelligence Score, HbA1c: Glycated hemoglobin, HDL: High density lipoprotein, HR: Heart rate, NO_2_: Nitrogen dioxide, SHBG: sex hormone binding globulin, PO4: phosphate, Vit D: Vitamin D

**Figure 7.**
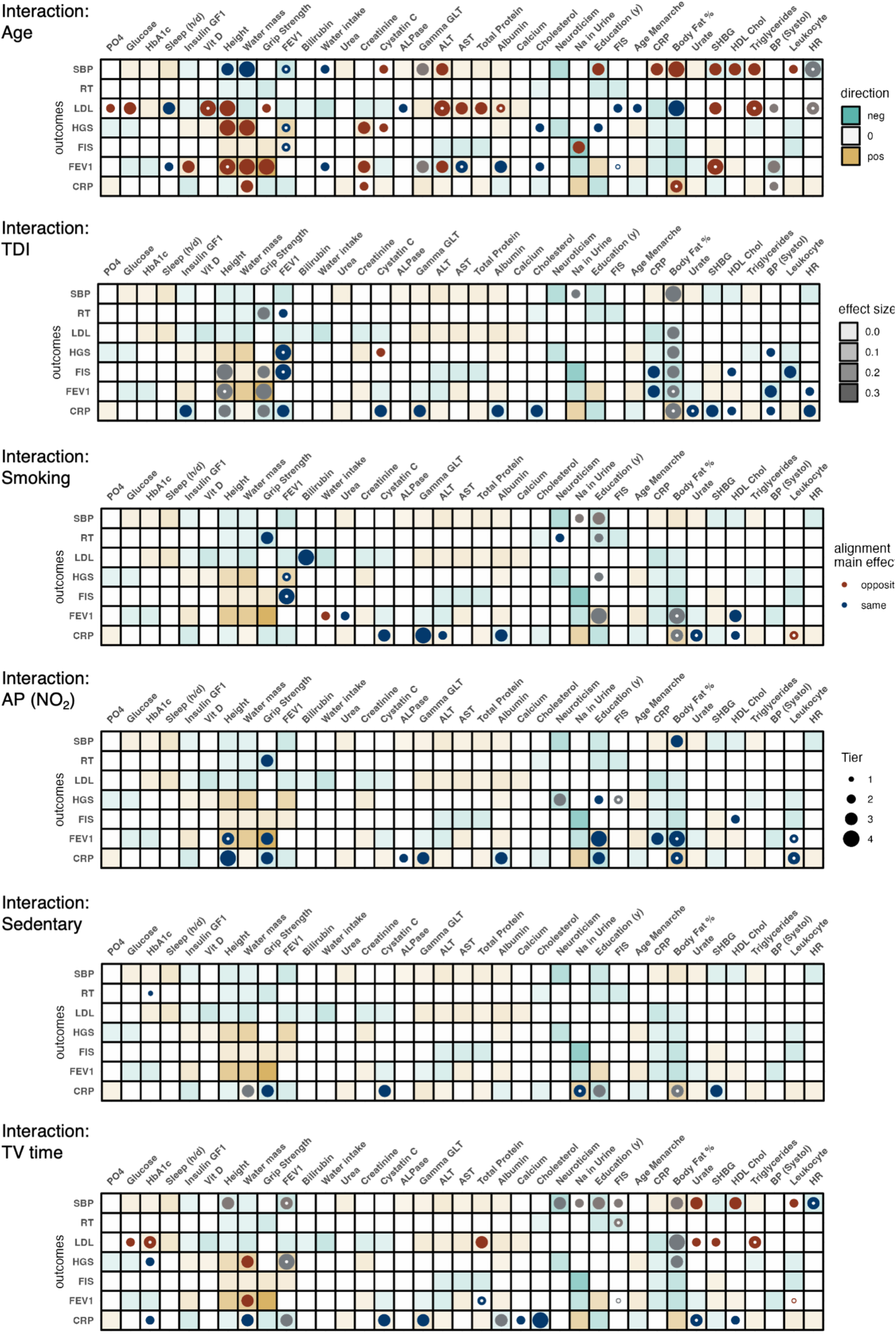
Results of the applied interaction analysis. Panels indicate interactions with each a different interaction parameter (age, socioeconomic deprivation (Townsend Deprivation Index, TDI), smoking, air pollution (NO_2_), sedentary behavior, and time spent watching TV (TV time). The dots indicate evidence for an interaction, with the size of the dots representing the tier score (1 = very little confidence, 2 = little confidence, 3 = some confidence, 4 = high confidence). The color of the dots indicates whether the interaction effect agrees with the direction of the main effect (dark blue) or not (red), i.e. whether the effect of the exposure on the outcome increases with higher levels of the environment (dark blue). The small white dots indicate when the corrected interaction estimate was considered due to the presence of a non-linear exposure-outcome relationship. Dots were shaded out if there was evidence for a strong causal effect of the exposure on the environment, as this may lead to biases in the interaction estimates. ALT: Alanine Aminotransferase, AST: Aspartate Aminotransferase, BP: Blood pressure, CRP: C-reactive Protein, FEV1: Forced Expiratory Volume within 1 second, FIS: Fluid Intelligence Score, HbA1c: Glycated hemoglobin, HDL: High density lipoprotein, HR: Heart rate, NO_2_: Nitrogen dioxide, SHBG: sex hormone binding globulin, PO4: phosphate, Vit D: Vitamin D

**Figure 8.**
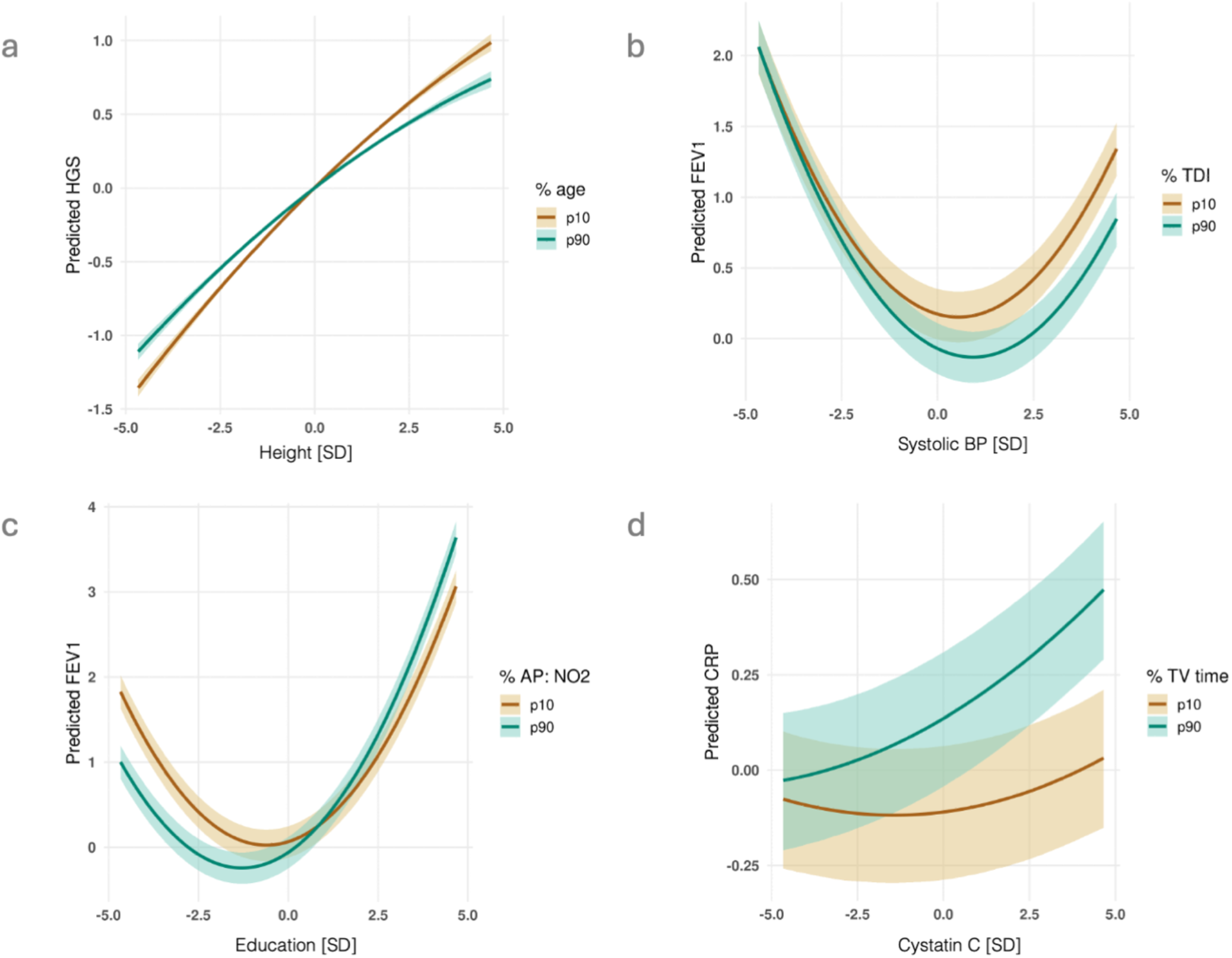
Plots of single interactions obtained using the full MR model. Selection of raw interactions obtained using the strict GRS. Units translate to the standard deviation (SD), and the confidence hulls indicate the 95R confidence interval. FEV1: Forced expiratory volume within 1 s, BP: blood pressure, p10: 10^th^ percentile, p90: 90st percentile, TDI: Townsend Deprivation Index, AP: Air pollution, CRP: C-reactive protein.

## Discussion

We present 2SLS-I, an approach to investigate how environmental variables modulate causal effects. Extensive simulations revealed accurate interaction estimates for almost all settings. Yet, we observed biased interaction estimates in presence of a non-linear exposure-outcome relationship and dependent environment and exposure, for which we provide a method to correct if indicated. Application of 2SLS-I to a range of health-related exposure- and outcome phenotypes revealed that all considered environments (age, socioeconomic deprivation (Townsend Deprivation Index, TDI), smoking, air pollution (nitrogen dioxide, NO_2_), physical inactivity (sedentary behaviour, PA: sed) and time spent watching TV (TV time)) modulate some causal relationships. We found that the strength of causal relationships tends to be attenuated in higher age. For example, our results indicate that age modulates the well-known relationship between height and hand grip strength (HGS). Whilst it is known that not only height (Abaraogu et al., 2017), but also age (e.g. Frederiksen et al., 2006) predicts HGS, it seems that the effects of age and height on HGS are not only additive, but in fact multiplicative. The tendency for causal effects to be attenuated in older individuals may be the result of the accumulation of other health-related factors which become more relevant the older a person grows.

Interestingly, we observed the opposite pattern for socioeconomic deprivation (Townsend Deprivation Index, TDI), where (mostly detrimental) causal effects seem to be intensified for people living in more deprived areas. For example, we found that the detrimental effect of systolic blood pressure (SBP) on forced expiratory volume (FEV1) was significantly intensified for people with lower socioeconomic status. Whilst there is some evidence for an association between hypertension and FEV1 (Miele et al., 2018), others argue that the effect is reverse, in fact, with lower FEV1 increasing blood pressure (Engström et al., 2001). Furthermore, there is also evidence that the negative association between blood pressure and FEV1 may result from confounding due to antihypertensive medication (Schnabel et al., 2011). Yet, causal inference methods point toward a negative effect of higher systolic blood pressure on FEV1 (Zekavat et al., 2021). Extending findings from Wheeler and colleagues (2005), who reported a positive association between socioeconomic status and FEV1, we provide evidence that the causal effect of SBP on FEV1 is exacerbated in response to socioeconomic deprivation.

For smoking, we observed that numerous causal effects were exacerbated in smokers in comparison to non-smokers. For example, we found that the causal effect of Gamma GLT on CRP is more pronounced in smokers than in non-smokers. Whilst it has long been known that Gamma GLT has a positive (i.e. increasing) effect on CRP (Lee et al., 2003), and studies found that smoking increases both, levels of Gamma GLT (Zhang et al., 2021) and CRP (O’Loughlin et al., 2008; Tracy et al., 1997), we provide evidence that the relationship between Gamma GLT and CRP is exacerbated in smokers in comparison to non-smokers.

Meanwhile, air pollution (NO_2_) mostly modulated effects on CRP and FEV1, whereby it exclusively intensified the causal effects. For example, the effect of education on FEV1 was intensified in response to living in an area with higher air pollution. Tabak and colleagues (2009) report a lower smoking adjusted FEV1 at baseline in people with a low educational level in comparison to people with a high educational level. Furthermore, it has long been known that short- and long-term exposure to air pollution (and NO_2_ in particular) is negatively associated with lung function (both FVC and FEV1) (Ackermann-Liebrich et al., 1997; Strassmann et al., 2021). Interestingly, multiple studies show some modulating effect of (parental) socioeconomic status on the association between air pollution and lung capacity (Cakmak et al., 2016; Wheeler, 2005). Yet, the effects seem to differ depending on participants’ sex and the definition (and potentially stratification) of the SES variable(s). A systematic comparison between varying definitions of education, potentially stratifying by sex, may contribute to a better understanding of the mechanisms through which education is protective against the detrimental effects of air pollution on lung function.

The effect modifying role of physical activity could only be assessed to a limited extent. As accelerometer data was only available in 67912 individuals after quality control, and physical inactivity seems to act as a collider in multiple relevant settings (e.g. the effect of body fat percentage on CRP), we only detected interactions with relatively low confidence (maximum tier score = 3). Importantly, physical inactivity mostly modulated effects on the outcome CRP (7 out of 8 interactions with a tier score > 0), of which all indicated that physical inactivity intensifies the (univariable) main effects. Given the well-known anti-inflammatory effect of physical activity, it is likely that physical inactivity exacerbates detrimental causal effects on CRP, which aligns with our observations. Nevertheless, it would be of great interest to investigate the effect modifying role of physical (in)activity more thoroughly in a larger sample.

The moderating effect of TV time turns out to be difficult to interpret. For example, we found that the positive effect of Cystatin C on CRP is significantly intensified in people who spend more time watching TV. Whilst there is evidence for an association between Cystatin C and CRP (Shlipak et al., 2005), it is worth noting that higher levels in Cystatin C have been found to be associated with higher age, higher triglycerides, lower HDL-Cholesterol and a range of inflammatory markers other than CRP, such as TNFα and Interleukin-6 (Luc et al., 2006). Meanwhile, TV time has been found to be positively associated with CRP even in children (Gabel et al., 2016) and after accounting for a range of relevant covariates such as waist circumference, physical activity, and dietary density (Gabel et al., 2016). As TV time is likely a function of many factors, such as age, general health, and socioeconomic status, and cannot be instrumented genetically, it remains difficult to investigate to what extent an intervention on TV time would reduce the health consequences associated with it.

Collectively, these results illustrate the relevance of environmental parameters as modulators of causal effects. Nevertheless, it is important to keep in mind limitations of the current study.

Firstly, whilst we tested our methods in a wide range of settings and simulated many (potential) sources of bias, from non-linear causal effects to linear and non-linear confounding, to the presence of level 1 interactions, simulations are typically incapable of accounting for the complexities of the real world. Whilst we are confident that 2SLS-I yields accurate effects in a wide range of settings, it is likely that there are specific circumstances where 2SLS-I fails to provide accurate findings.

Second, the detection of interactions has low statistical power. To address this, we replicated our analysis using an extended GRS (GRS_ext_), which may violate MR assumptions (the weaker SNPs correlate with the exposure, the more likely that they act indirectly). To account for this circumstance, we ensured that the results from the GRS_ext_ only contributed to an increased tier score if the effect estimate agreed with the respective estimate of the strict GRS. Furthermore, we considered interactions as “significant ” if they reached a p-value < 0.001, which allowed us to detect more potential interactions, but also increases the false discovery rate.

Another way to increase power is by carefully choosing when to apply the corrected interaction estimates. Since the 2SLS-I-corr effects have larger variance than the uncorrected counterpart, they should primarily be used when there is evidence for a non-linear causal effect. The replication of the present findings in an additional cohort would be necessary to confidently consider them as valid, in particular those with a tier score < 4. Such interactions, however, may be population-specific, which renders replication particularly difficult. Furthermore, despite making a considerable effort to account for the reduced power inherent to interaction analysis (and Mendelian Randomization), the absence of a significant interaction may still be the result of lacking power and does not translate to a proof of inexistence, in particular for phenotypes with relatively small sample size and high variance of noise, such as physical inactivity. Yet, thanks to the ever-increasing sample size of available biobanks, we are confident that the reduced power of interaction analyses will become a smaller challenge in the future. If this is the case, 2SLS-I would even allow for the investigation of environment-dependent non-linear effects (e.g. *X*^2^ · *E*), which may be of relevance as we observed multiple settings with evidence for non-linear causal effects. Still, it has to be noted that we modelled only quadratic X-to-Y effects and our proposed correction is suboptimal for more complex non-linear X-Y relationships.

Third, due to the far-reaching effects of many environmental parameters, some interactions are difficult to interpret. A detected exposure-environment interaction may arise only due to a true interaction between the exposure and another (correlated) environment. Whilst understanding that an environmental variable modulates a causal effect is of great relevance, we note that the present analysis could (and should) be extended for almost all detected interactions to obtain a detailed understanding of the complex interplay between variables, in particular for those related to SES.

Fourth, detected interactions may be scale specific and not persist when modelling a transformed version of the outcome (e.g. *log(Y)*). This is a general weakness of all types of interaction analyses. Nevertheless, we replicated our analysis by inverse-rank normal transforming the outcome phenotypes and found that most interactions yield comparable effects independent of the outcome standardization. Yet, in particular for CRP we observed multiple settings where the effect seems to depend on the standardization of the outcome phenotype due to the strong right-skewedness of CRP.

Fifth, one has to carefully exclude the possibility that a tested environment is a collider of the exposure-outcome relationship, because it can lead to biased interaction estimates. If such situation occurs, it is safer to regress out the exposure from the environmental variable and consider the residual trait as the new tested environment. Similarly, analyses should assess the presence and relevance of *G* · *E* effects on X as they violate our assumption that the genetic effects on the exposure are environment-invariant.

Sixth, the full overlap between samples from which the SNP-exposure and SNP-outcome effects are estimated can introduce bias in the interaction estimates, which requires further work to account for. However, based on the impact of sample overlap on the causal main effect, (Mounier *S* Kutalik, 2023), it is likely to play a minor role for interactions too.

Furthermore, we did not aim at validating the non-linear effects observed using 2SLS-I. Although we obtained accurate non-linear estimates for all simulated settings, future projects could aim at extending the simulation settings to challenge the accuracy of 2SLS-I for non-linear effects or perform negative control experiments to investigate if the non-linear effects of 2SLS-I are robust across a wide range of settings.

Finally, the present analysis was restricted to individuals of White British ancestry. As many environmental parameters, such as SES, vary between different ancestries, it would be of great relevance to extend the present analyses to diverse ancestral groups.

In conclusion, we present 2SLS-I, a method to investigate how environmental variables modulate causal effects, even in presence of sources of bias, such as non-linear effects of the exposure on the outcome. We demonstrate the reliability of 2SLS-I in a wide range of simulation settings. Finally, we provide evidence that it is relevant to consider the modulating effects of environmental variables such as age and SES when examining the causal effect of classical epidemiological risk factors, which is a step towards precision medicine.

## Data Availability

The data that support the findings of this study are available from the UK Biobank (UKBB), but restrictions apply to the availability of these data, which were used under license for the current study. Access to the UKBB can be requested through a standard protocol (https://www.ukbiobank.ac.uk/enable-your-research/apply-for-access). Summary statistics were accessed from Neale and colleagues (2017), which are publicly available (http://www.nealelab.is/uk-biobank), just as the weights to calculate the extended genetic risk score as in Prive et al. (2022) (https://www.pgscatalog.org/publication/PGP000263/).

http://www.nealelab.is/uk-biobank

https://www.pgscatalog.org/publication/PGP000263/

## Data availability

The data that support the findings of this study are available from the UK Biobank (UKBB), but restrictions apply to the availability of these data, which were used under license for the current study. Access to the UKBB can be requested through a standard protocol (https://www.ukbiobank.ac.uk/enable-your-research/apply-for-access). Summary statistics were accessed from Neale and colleagues (2017), which are publicly available (http://www.nealelab.is/uk-biobank), just as the weights to calculate the extended genetic risk score as in Privé et al. (2022) (https://www.pgscatalog.org/publication/PGP000263/).

## Code availability

Scripts used to perform the analyses are available at https://github.com/leonakn/2SLS-I.

## Additional information

### Ethical approval

The UK Biobank has approval from the North West Multi-centre Research Ethics Committee (MREC). The data was accessed through application number 16389.

## Supplementary Material

### Sensitivity Analyses

Multiple sensitivity analyses were performed to account for potential biases in our effects. Amongst them are the effect of level 1 interactions, meaning an interaction between the GRS and the environment on the exposure X, the potential for the environment to act as a collider between the exposure and the outcome, the risk for the interactions to depend on the standardization or transformation of the outcome variable and the potential for the detected interactions to be method-specific.

### Level 1 interactions

**Supplementary Figure 1:**
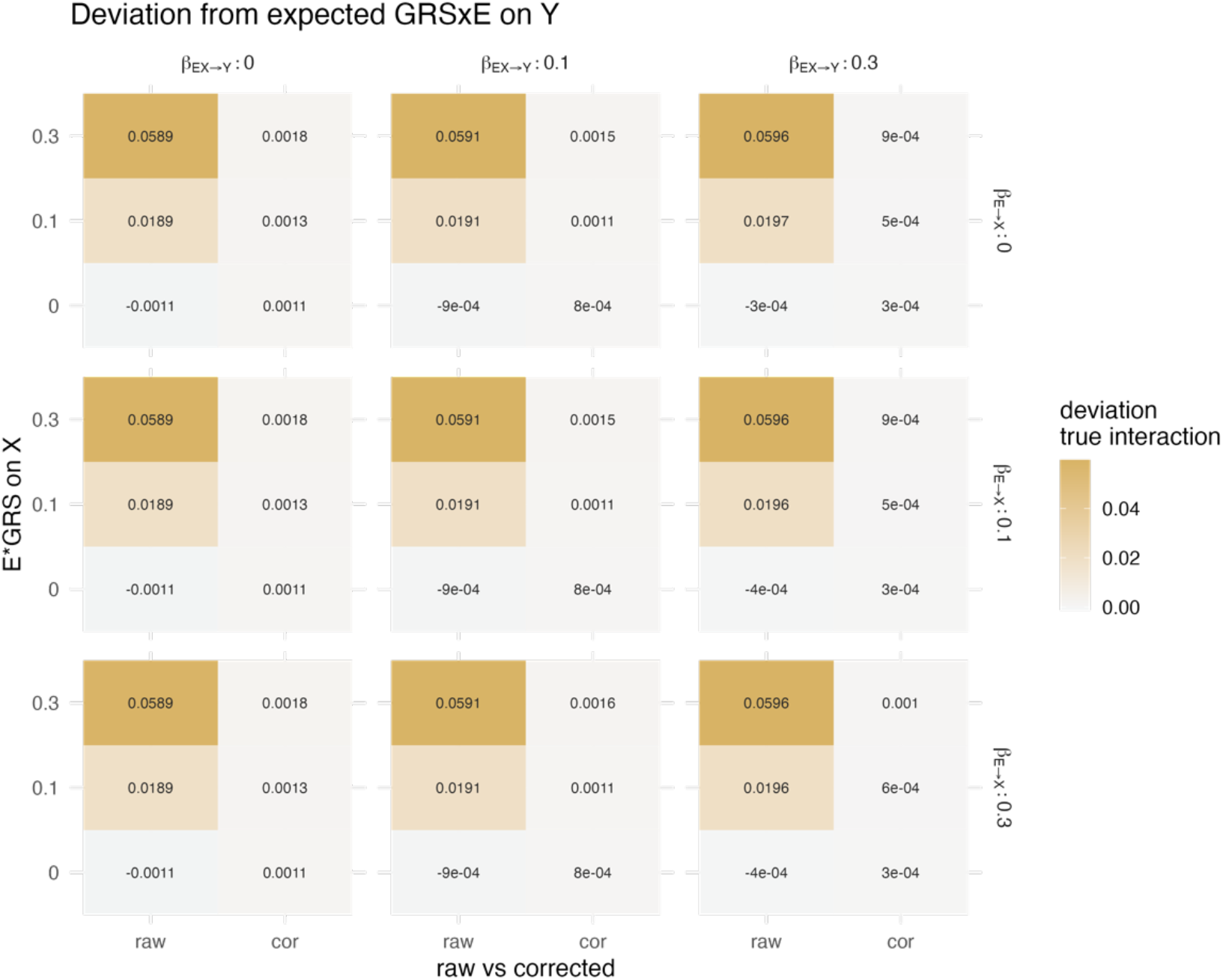
Bias in interaction effect in response to varying level 1 interactions if the simulated effect of the exposure on the outcome is 0.2. We observe that the bias translates to 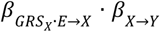 and can be corrected for by subtracting the according term from the interaction estimate, which attenuated the bias in interaction estimates to zero.

Assuming the exposure X is defined as follows:

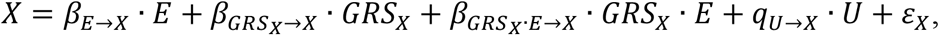

while the outcome Y is – amongst other things – a function of X, and 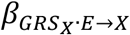 deviates from zero, we will overestimate the effect of 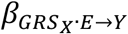 by 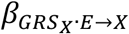· *β*_*X*→*Y*_. We validated this in simulation settings (*Supplementary Figure 1*). We investigated to what extent the presence of level 1 interactions may affect our interaction estimates from the application study. For 160 interactions with a tier score > 0, there was none where correction for level 1 interaction would lead to a change in the sign of the interaction estimate. Furthermore, there were only 5 settings where the absolute ratio between the obtained interaction estimate and the correction was smaller than 5, namely the interaction between Body fat percentage and Nitrogen dioxide on CRP, the interactions between water mass and TV time on forced expiratory volume and hand grip strength, the interaction between CRP and age on systolic blood pressure and finally the interaction between height and socioeconomic deprivation on forced expiratory volume.

### Exposure on environment effects

For some settings, we observed that there was a causal effect of the exposure on the environment. For these settings, we obtained the (observational, as the environment often is not meaningfully genetically instrumentable) effect of the environment on the exposure. We obtained the ratio 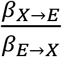 and the according 95% confidence intervals. If the lower absolute confidence interval of the 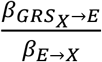 ratio was larger than 0.5, we considered the interaction as potentially biased and shaded it out in the according plots. For example, for body fat percentage we found a relatively strong effect on SES, TV time, and sedentary behavior, relative to the reverse effect. Noteworthily, we also found evidence for some causal effects on age, namely of heart rate, Gamma GLT, and SBP. Those likely result from sample bias, meaning that people with genetically high systolic blood pressure do not feel well enough to participate in such a study at an age where their peers still are able to participate.

### Inverse rank normal transformed outcome phenotypes

We replicated our analyses whereby we inverse rank normal transformed the outcome phenotypes (which by default were z-standardized). For the majority of settings (136 out of 182 interactions with a tier score > 0, 74.7%), the IRNT outcome phenotypes yielded interaction results that agreed well (tier > 0, agreement in direction) with the z-standardized outcome phenotypes. It is worth nothing, though, that we observed in 44 (24.2%) of all interactions with a tier score > 0, that the result could not be replicated when the outcome was IRN-transformed (tier score for IRN-transformed outcome = 0). There were two settings where both, the interaction on the IRN-transformed outcome and the interaction on the z-standardized outcome yielded a tier score > 0 but the direction of the effect did not agree, namely the interation between FEV1 and age on HGS and the interaction between Body fat % and age on CRP. Finally, there were 55 interactions which only yielded a tier score > 0 when the outcome was IRN-transformed, but not when it was z standardized. The majority of interactions that deviated depending on the preprocessing of the outcome phenotype were interactions affecting CRP (64 of 101 settings), RT (12 of 101 settings), and FEV1 (11 of 101 settings). This finding is little surprising given the heavy right skewing of the raw (and thus z-standardized) CRP phenotype, indicating that some of the interactions may be driven by the extreme values on the upper end of the scale. As these values may be of distinctive mechanistic relevance, it depends on the specific research question whether it is more accurate to consider the results obtained using the z-standardized or the IRN-transformed outcome phenotype.

**Supplementary Figure 2.**
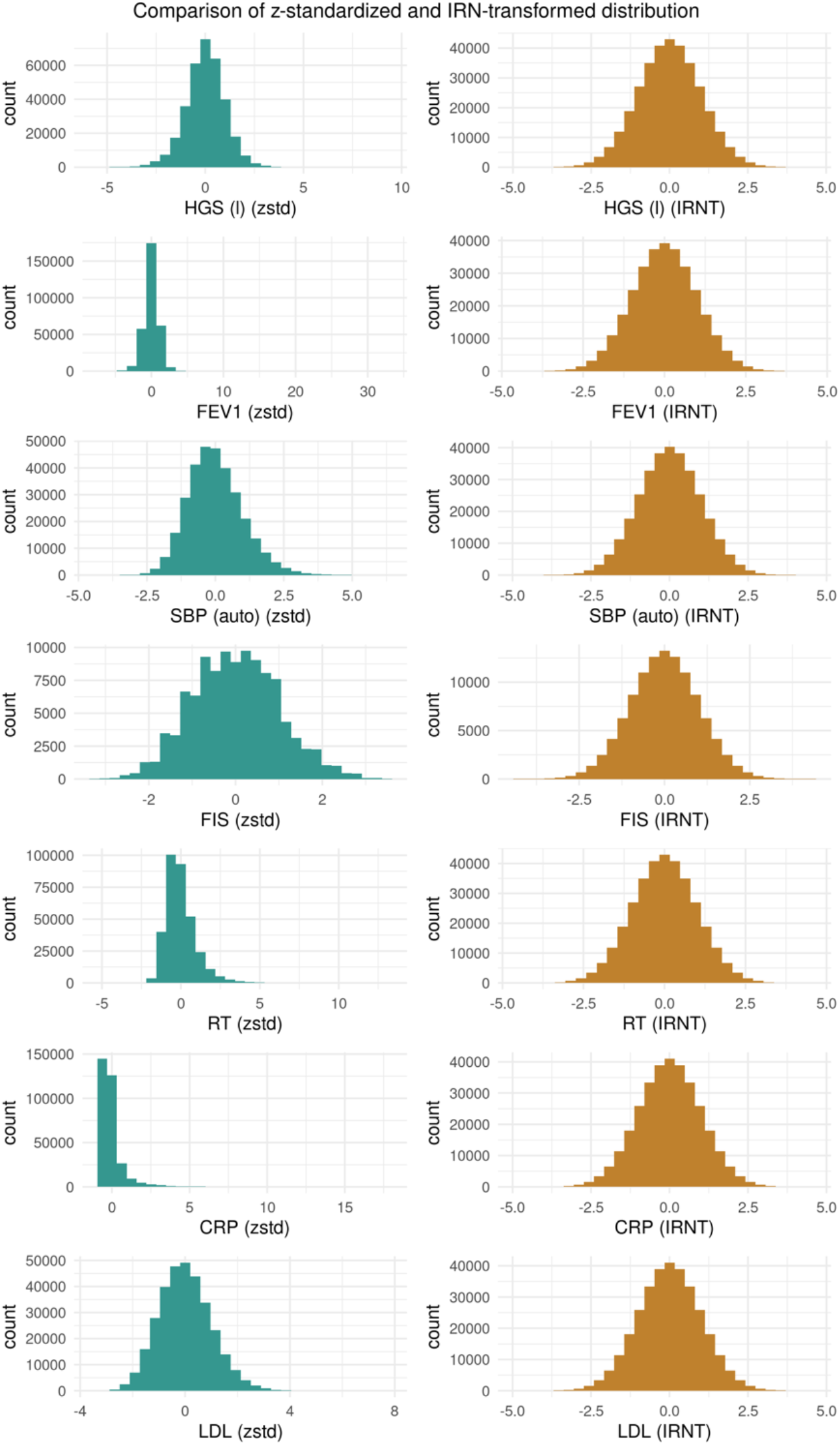
Density plots of the z-standardized (zstd, blue, left) vs. inverse rank normal transformed (IRNT, right) outcome phenotypes. HGS (l): hand grip strength (left), FEV1: forced expiratory volume within 1 second, SBP (auto): systolic blood pressure, automated reading, FIS: fluid intelligence score, RT: reaction time at pattern matching task, CRP: C-reactive protein, LDL: Low density Lipoprotein.

**Supplementary Figure 3.**
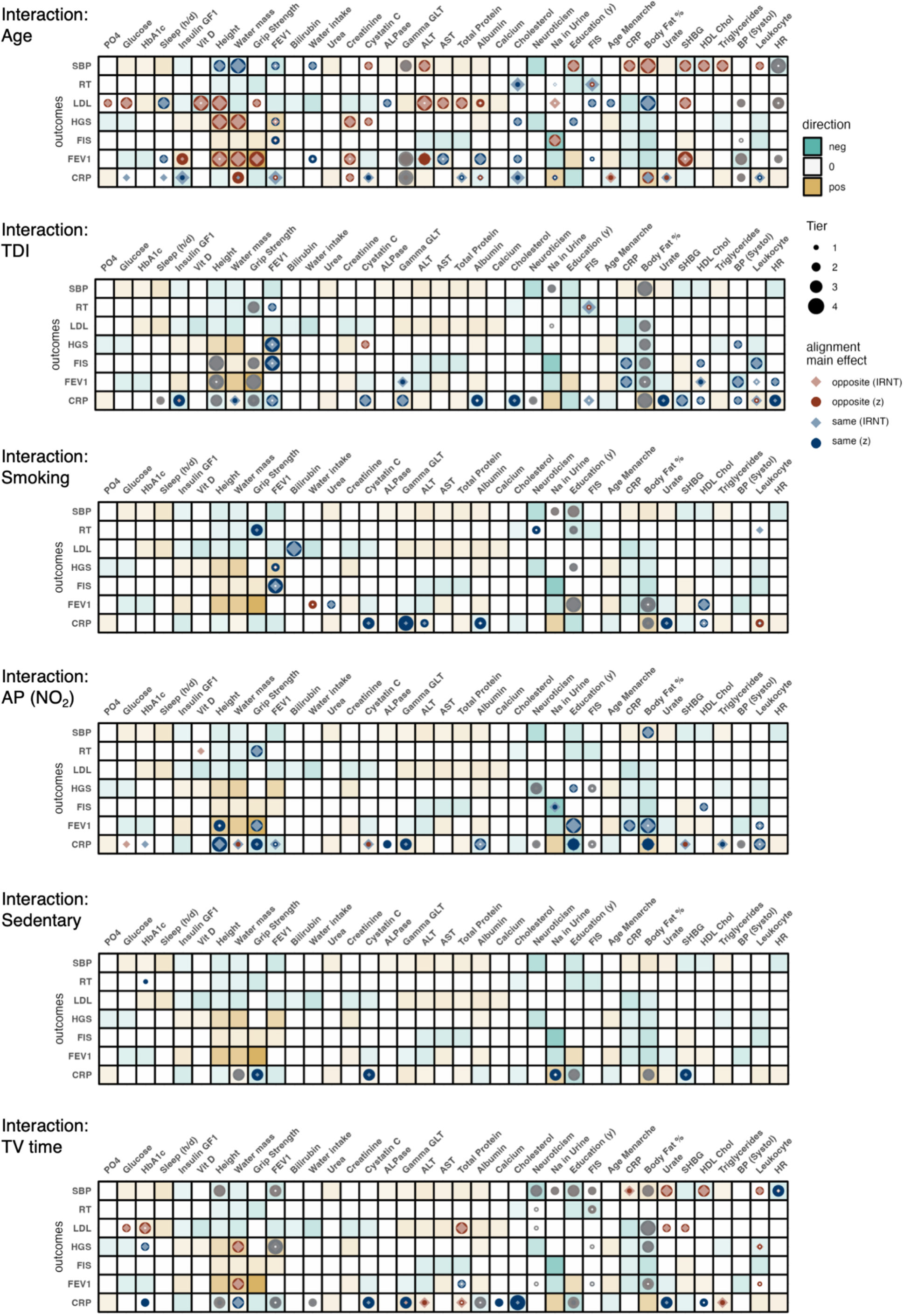
Replication of findings when outcome phenotype is not z-standardized but inverse-rank normal transformed (IRNT). The color of the tiles represents the direction of the main effect if there was evidence for a main effect. The round shapes represent the interactions with a tier score > 0 obtained when the outcome was z-standardized, whereby the color indicates the direction of the interaction relative to the main effect. The rhombuses represent the interactions with tier > 0 obtained when the outcome phenotype was inverse rank normal transformed, whereby the color indicates the direction of the interaction relative to the main effect. The size represents the tier score, with larger shapes indicating higher confidence. The small white dot indicates if an interaction was corrected due to evidence for a quadratic effect of the exposure on the outcome and an effect of the environment on the exposure. Finally, interactions are shaded out if there was evidence for a stronger effect of the exposure on the environment than vice versa, as this likely leads to biased interaction estimates. AP: Air pollution, ALT: Alanine Aminotransferase, AST: Aspartate Aminotransferase, BP: Blood pressure, CRP: C-reactive Protein, FEV1: Forced Expiratory Volume within 1 second, FIS: Fluid Intelligence Score, HbA1c: Glycated hemoglobin, HDL: High density lipoprotein, HR: Heart rate, NO_2_: Nitrogen dioxide, SHBG: sex hormone binding globulin, PO4: phosphate, TDI: Townsend deprivation index, Vit D: Vitamin D

### Replication analysis

To validate the present interaction effects, we assessed the same settings in an extended doubly ranked stratification (Tian et al., 2023) framework (DRS). Whilst the doubly ranked stratification method was developed to investigate non-linear exposure-outcome relationships, it can be adapted to allow for investigating interactions by stratifying by an environmental variable (instead of the exposure value) adjusted by the exposure’s instrument and regressing the obtained MR estimates on the predefined strata midpoint value. Applying DRS to the same phenotype combination as 2SLS-I, we found substantial agreement between the two methods. In 136 (85 %) of all interactions with a tier score > 0, the direction of DRS aligned. In 30 (18.75%) of all interactions with a tier score > 0, the doubly ranked method agreed in sign and significance. Vice versa, out of 50 settings where DRS yielded significance at p < 0.001, 2SLS-I yielded a tier score > 0 in 30 (60 %).There was no setting where both, the DRS (at p < 0.001) and 2SLS-I (tier > 0) yielded significant effects which did not agree in sign. In summary, DRS and 2SLS-I yield comparable results (*Supplementary Figure 4)* Whilst this contributes to the confidence in the results obtained with 2SLS-I, it should be noted that there is a potential for both, 2SLS-I and the DRS yielding false-positive (e.g. if the environment acts as collider between the exposure and the outcome) or false-negative (e.g. due to lacking power) results.

**Supplementary Figure 4.**
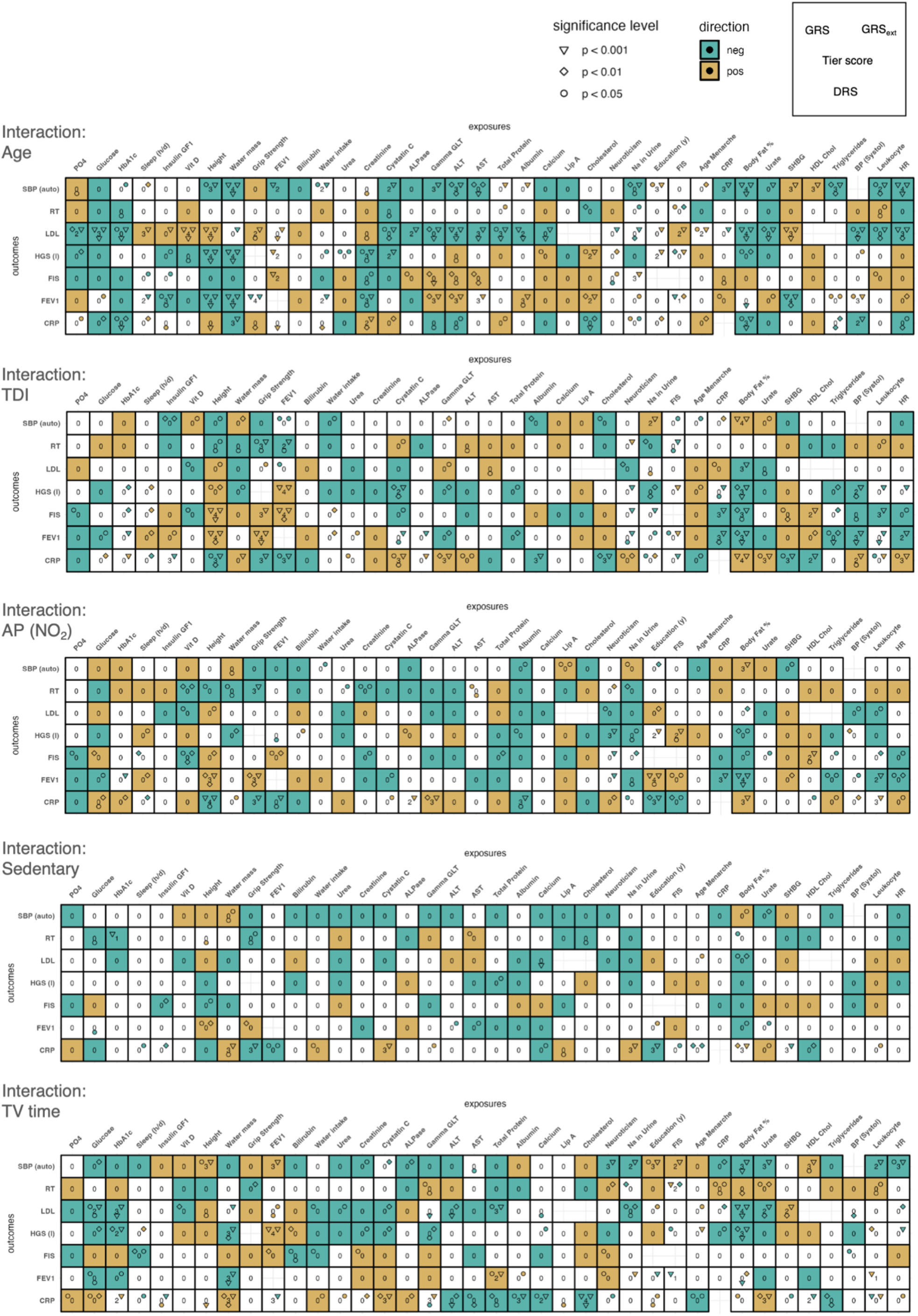
All interaction effects from 2SLS-I were replicated using a doubly ranked stratification (DRS) method. The figure summarizes the effect agreement between 2SLS-I and DRS. Tiles are filled if all three estimates (strict GRS of 2SLS-I, extended GRS of 2SLS-I, and DRS) agree in the direction of the effect, with the fill color indicating the direction of the interaction. For each estimate that yielded significance at p < 0.05, a shape is plotted to indicate the level of significance: a circle (p < 0.05), a rhombus (p < 0.01) or a triangle (p < 0.001). The position of the according shape within each tile indicates which method has yielded the according significance level, with the strict GRS of 2SLS-I being on the top left, the extended GRS of 2SLS-I on the top right and the DRS at the bottom. The color of each of these shapes indicates the direction of the effect. Finally, the number in the middle of each tile relates to the obtained tier score, i.e. the confidence rating obtained using 2SLS-I.

## References

Abaraogu, U. O., Ezema, C. I., Ofodile, U. N., & Igwe, S. E. (2017). Association of grip strength with anthropometric measures: Height, forearm diameter, and middle finger length in young adults. Polish Annals of Medicine, 24(2), 153–157. 10.1016/j.poamed.2016.11.008

Ackermann-Liebrich, U., Leuenberger, P., Schwartz, J., Schindler, C., Monn, C., Bolognini, G., Bongard, J. P., Brändli, O., Domenighetti, G., Elsasser, S., Grize, L., Karrer, W., Keller, R., Keller-Wossidlo, H., Künzli, N., Martin, B. W., Medici, T. C., Perruchoud, A. P., Schöni, M. H., … Zemp, E. (1997). Lung function and long term exposure to air pollutants in Switzerland. Study on Air Pollution and Lung Diseases in Adults (SAPALDIA) Team. American Journal of Respiratory and Critical Care Medicine, 155(1), 122–129. 10.1164/ajrccm.155.1.9001300

Berrington de Gonzalez, A., Hartge, P., Cerhan, J. R., Flint, A. J., Hannan, L., MacInnis, R. J., Moore, S. C., Tobias, G. S., Anton-Culver, H., Freeman, L. B., Beeson, W. L., Clipp, S. L., English, D. R., Folsom, A. R., Freedman, D. M., Giles, G., Hakansson, N., Henderson, K. D., Hoffman-Bolton, J., … Thun, M. J. (2010). Body-Mass Index and Mortality among 1.46 Million White Adults. New England Journal of Medicine, 363(23), 2211–2219. 10.1056/NEJMoa1000367

Cakmak, S., Hebbern, C., Cakmak, J. D., & Vanos, J. (2016). The modifying effect of socioeconomic status on the relationship between traffic, air pollution and respiratory health in elementary schoolchildren. Journal of Environmental Management, 177, 1–8. 10.1016/j.jenvman.2016.03.051

Collister, J. A., Liu, X., & Clifton, L. (2022). Calculating Polygenic Risk Scores (PRS) in UK Biobank: A Practical Guide for Epidemiologists. Frontiers in Genetics, 13, 818574. 10.3389/fgene.2022.818574

Cortina, J. M. (1993). Interaction, Nonlinearity, and Multicollinearity: Implications for Mu/tip/e Regression. 19(4), 915–922.

Engström, G., Wollmer, P., Valind, S., Hedblad, B., & Janzon, L. (2001). Blood pressure increase between 55 and 68 years of age is inversely related to lung function: Longitudinal results from the cohort study ‘Men born in 1914’: Journal of Hypertension, 19(7), 1203–1208. 10.1097/00004872-200107000-00004

Frederiksen, H., Hjelmborg, J., Mortensen, J., Mcgue, M., Vaupel, J., & Christensen, K. (2006). Age Trajectories of Grip Strength: Cross-Sectional and Longitudinal Data Among 8,342 Danes Aged 46 to 102. Annals of Epidemiology, 16(7), 554–562. 10.1016/j.annepidem.2005.10.006

Gabel, L., Ridgers, N. D., Della Gatta, P. A., Arundell, L., Cerin, E., Robinson, S., Daly, R. M., Dunstan, D. W., & Salmon, J. (2016). Associations of sedentary time patterns and TV viewing time with inflammatory and endothelial function biomarkers in children. Pediatric Obesity, 11(3), 194–201. 10.1111/ijpo.12045

Lawlor, D. A., Harbord, R. M., Sterne, J. A. C., Timpson, N., & Davey Smith, G. (2008). Mendelian randomization: Using genes as instruments for making causal inferences in epidemiology. Statistics in Medicine, 27(8), 1133–1163. 10.1002/sim.3034

Lee, D.-H., Jacobs, D. R., Gross, M., Kiefe, C. I., Roseman, J., Lewis, C. E., & Steffes, M. (2003). Glutamyltransferase Is a Predictor of Incident Diabetes and Hypertension: TheCoronary Artery Risk Development in Young Adults (CARDIA) Study. Clinical Chemistry, 8.

Liu, Y., Elsworth, B., Erola, P., Haberland, V., Hemani, G., Lyon, M., Zheng, J., Lloyd, O., Vabistsevits, M., & Gaunt, T. R. (2021). EpiGraphDB: A database and data mining platform for health data science. Bioinformatics, 37(9), 1304–1311. 10.1093/bioinformatics/btaa961

Luc, G., Bard, J.-M., Lesueur, C., Arveiler, D., Evans, A., Amouyel, P., Ferrieres, J., Juhan-Vague, I., Fruchart, J.-C., & Ducimetiere, P. (2006). Plasma cystatin-C and development of coronary heart disease: The PRIME Study. Atherosclerosis, 185(2), 375–380. 10.1016/j.atherosclerosis.2005.06.017

Matuschek, H., & Kliegl, R. (2018). On the ambiguity of interaction and nonlinear main effects in a regime of dependent covariates. Behavior Research Methods, 50(5), 1882–1894. 10.3758/s13428-017-0956-9

McCaw, Z. R., Lane, J. M., Saxena, R., Redline, S., & Lin, X. (2020). Operating characteristics of the rank-based inverse normal transformation for quantitative trait analysis in genome-wide association studies. Biometrics, 76(4), 1262–1272. 10.1111/biom.13214

Miele, C. H., Grigsby, M. R., Siddharthan, T., Gilman, R. H., Miranda, J. J., Bernabe-Ortiz, A., Wise, R. A., & Checkley, W. (2018). Environmental exposures and systemic hypertension are risk factors for decline in lung function. Thorax, 73(12), 1120–1127. 10.1136/thoraxjnl-2017-210477

Mölder, F., Jablonski, K. P., Letcher, B., Hall, M. B., Tomkins-Tinch, C. H., Sochat, V., Forster, J., Lee, S., Twardziok, S. O., Kanitz, A., Wilm, A., Holtgrewe, M., Rahmann, S., Nahnsen, S., & Köster, J. (2021). Sustainable data analysis with Snakemake. F1000Research, 10, 33. 10.12688/f1000research.29032.2

Mounier, N., & Kutalik, Z. (2023). Bias correction for inverse variance weighting Mendelian randomization. Genetic Epidemiology, 47(4), 314–331. 10.1002/gepi.22522

Neale. (2017, September 20). Rapid GWAS of thousands of phenotypes for 337,000 samples in the UK Biobank. http://www.nealelab.is/blog/2017/7/19/rapid-gwas-of-thousands-of-phenotypes-for-337000-samples-in-the-uk-biobank

O’Loughlin, J., Lambert, M., Karp, I., McGrath, J., Gray-Donald, K., Barnett, T., Delvin, E., Levy, E., & Paradis, G. (2008). Association between cigarette smoking and C-reactive protein in a representative, population-based sample of adolescents. Nicotine & Tobacco Research, 10(3), 525–532. 10.1080/14622200801901997

Privé, F., Aschard, H., Carmi, S., Folkersen, L., Hoggart, C., O’Reilly, P. F., & Vilhjálmsson, B. J. (2022). Portability of 245 polygenic scores when derived from the UK Biobank and applied to 9 ancestry groups from the same cohort. The American Journal of Human Genetics, 109(1), 12–23. 10.1016/j.ajhg.2021.11.008

R Core Team. (2022). R: A language and environment for statistical computing. (4.2.1) [Computer software]. R Foundation for Statistical Computing.

Richardson, T. G., Sanderson, E., Elsworth, B., Tilling, K., & Davey Smith, G. (2020). Use of genetic variation to separate the effects of early and later life adiposity on disease risk: Mendelian randomisation study. BMJ, m1203. 10.1136/bmj.m1203

Sanderson, E., Glymour, M. M., Holmes, M. V., Kang, H., Morrison, J., Munafò, M. R., Palmer, T., Schooling, C. M., Wallace, C., Zhao, Q., & Davey Smith, G. (2022). Mendelian randomization. Nature Reviews Methods Primers, 2(1), 6. 10.1038/s43586-021-00092-5

Schnabel, E., Nowak, D., Brasche, S., Wichmann, H.-E., & Heinrich, J. (2011). Association between lung function, hypertension and blood pressure medication. Respiratory Medicine, 105(5), 727–733. 10.1016/j.rmed.2010.12.023

Shlipak, M. G., Katz, R., Cushman, M., Sarnak, M. J., Stehman-Breen, C., Psaty, B. M., Siscovick, D., Tracy, R. P., Newman, A., & Fried, L. (2005). Cystatin-C and inflammatory markers in the ambulatory elderly. The American Journal of Medicine, 118(12), 1416.e25-1416.e31. 10.1016/j.amjmed.2005.07.060

Strassmann, A., De Hoogh, K., Röösli, M., Haile, S. R., Turk, A., Bopp, M., Puhan, M. A., & for the Swiss National Cohort Study Group. (2021). NO2 and PM2.5 Exposures and Lung Function in Swiss Adults: Estimated Effects of Short-Term Exposures and Long-Term Exposures with and without Adjustment for Short-Term Deviations. Environmental Health Perspectives, 129(1), 017009. 10.1289/EHP7529

Sudlow, C., Gallacher, J., Allen, N., Beral, V., Burton, P., Danesh, J., Downey, P., Elliott, P., Green, J., Landray, M., Liu, B., Matthews, P., Ong, G., Pell, J., Silman, A., Young, A., Sprosen, T., Peakman, T., & Collins, R. (2015). UK Biobank: An Open Access Resource for Identifying the Causes of a Wide Range of Complex Diseases of Middle and Old Age. PLOS Medicine, 12(3), e1001779. 10.1371/journal.pmed.1001779

Sulc, J., Sjaarda, J., & Kutalik, Z. (2021). Polynomial Mendelian Randomization reveals widespread non-linear causal effects in the UK Biobank [Preprint]. Genetics. 10.1101/2021.12.08.471751

Sun, Y.-Q., Burgess, S., Staley, J. R., Wood, A. M., Bell, S., Kaptoge, S. K., Guo, Q., Bolton, T. R., Mason, A. M., Butterworth, A. S., Di Angelantonio, E., Vie, G. Å., Bjørngaard, J. H., Kinge, J. M., Chen, Y., & Mai, X.-M. (2019). Body mass index and all cause mortality in HUNT and UK Biobank studies: Linear and non-linear mendelian randomisation analyses. BMJ, l1042. 10.1136/bmj.l1042

Tabak, C., Spijkerman, A. M. W., Verschuren, W. M. M., & Smit, H. A. (2009). Does educational level influence lung function decline (Doetinchem Cohort Study)? European Respiratory Journal, 34(4), 940–947. 10.1183/09031936.00111608

Tian, H., Mason, A. M., Liu, C., & Burgess, S. (2023). Relaxing parametric assumptions for non-linear Mendelian randomization using a doubly-ranked stratification method. PLOS Genetics, 19(6), e1010823. 10.1371/journal.pgen.1010823

Tracy, R. P., Psaty, B. M., Macy, E., Bovill, E. G., Cushman, M., Cornell, E. S., & Kuller, L. H. (1997). Lifetime Smoking Exposure Affects the Association of C-Reactive Protein with Cardiovascular Disease Risk Factors and Subclinical Disease in Healthy Elderly Subjects. Arteriosclerosis, Thrombosis, and Vascular Biology, 17(10), 2167–2176. 10.1161/01.ATV.17.10.2167

Wheeler, B. W. (2005). Environmental equity, air quality, socioeconomic status, and respiratory health: A linkage analysis of routine data from the Health Survey for England. Journal of Epidemiology & Community Health, 59(11), 948–954. 10.1136/jech.2005.036418

Zekavat, S. M., Honigberg, M., Pirruccello, J. P., Kohli, P., Karlson, E. W., Newton-Cheh, C., Zhao, H., & Natarajan, P. (2021). Elevated Blood Pressure Increases Pneumonia Risk: Epidemiological Association and Mendelian Randomization in the UK Biobank. Med, 2(2), 137-148.e4. 10.1016/j.medj.2020.11.001

Zhang, Z., Ma, L., Geng, H., & Bian, Y. (2021). Effects of Smoking, and Drinking on Serum Gamma-Glutamyl Transferase Levels Using Physical Examination Data: A Cross-Sectional Study in Northwest China. International Journal of General Medicine, Volume 14, 1301–1309. 10.2147/IJGM.S301900

